# Total-body Dynamic Imaging and Kinetic Modeling of ^18^F-AraG in Healthy Individuals and a Non-Small Cell Lung Cancer Patient Undergoing Anti-PD-1 Immunotherapy

**DOI:** 10.1101/2023.09.22.23295860

**Authors:** Negar Omidvari, Jelena Levi, Yasser G Abdelhafez, Yiran Wang, Lorenzo Nardo, Megan E Daly, Guobao Wang, Simon R Cherry

## Abstract

Immunotherapies, especially the checkpoint inhibitors such as anti-PD-1 antibodies, have transformed cancer treatment by enhancing immune system’s capability to target and kill cancer cells. However, predicting immunotherapy response remains challenging. ^18^F-AraG is a molecular imaging tracer targeting activated T cells, which may facilitate therapy response assessment by non-invasive quantification of immune cell activity within tumor microenvironment and elsewhere in the body. The aim of this study was to obtain preliminary data on total-body pharmacokinetics of ^18^F-AraG, as a potential quantitative biomarker for immune response evaluation.

**Methods:** The study consisted of 90-min total-body dynamic scans of four healthy subjects and one non-small cell lung cancer (NSCLC) patient, scanned before and after anti-PD-1 immunotherapy. Compartmental modeling with Akaike information criterion model selection were employed to analyze tracer kinetics in various organs. Additionally, seven sub-regions of the primary lung tumor and four mediastinal lymph nodes were analyzed. Practical identifiability analysis was performed to assess reliability of kinetic parameter estimation. Correlations of SUVmean, SUVR (tissue-to-blood ratio), and Logan plot slope (*K*_*Logan*_) with total volume-of-distribution (*V_T_*) were calculated to identify potential surrogates for kinetic modeling.

**Results:** Strong correlations were observed between *K*_*Logan*_ and SUVR values with *V_T_*, suggesting that they can be used as promising surrogates for *V_T_*, especially in organs with low blood-volume fraction. Moreover, the practical identifiability analysis suggests that the dynamic ^18^F-AraG PET scans could potentially be shortened to 60 minutes, while maintaining quantification accuracy for all organs-of-interest. The study suggests that although ^18^F-AraG SUV images can provide insights on immune cell distribution, kinetic modeling or graphical analysis methods may be required for accurate quantification of immune response post-therapy. While SUVmean showed variable changes in different sub-regions of the tumor post-therapy, the SUVR, *K*_*Logan*_, and *V_T_* showed consistent increasing trends in all analyzed sub-regions of the tumor with high practical identifiability.

**Conclusion:** Our findings highlight the promise of ^18^F-AraG dynamic imaging as a non-invasive biomarker for quantifying the immune response to immunotherapy in cancer patients. The promising total-body kinetic modeling results also suggest potentially wider applications of the tracer in investigating the role of T cells in the immunopathogenesis of diseases.

## INTRODUCTION

Immunotherapy has revolutionized cancer treatment by enhancing the capability of patient’s immune system to target and eliminate cancer cells. One of the most widely used immunotherapy classes are immune checkpoint inhibitors targeting the programmed cell death protein 1 (PD-1)/programmed death-ligand 1 (PD-L1) axis. These antibodies disrupt the inhibitory signals between cancer cells and T cells to reinvigorate the immune response. Despite the remarkable clinical successes of anti-PD-1/anti-PD-L1 immunotherapies, not all patients respond equally to treatment, highlighting the need for early predictive biomarkers to identify potential responders. Predicting immunotherapy response is challenging due to the complexity of the tumor microenvironment and the systemic nature of the immune response, particularly involving the lymph nodes (LNs). The dynamic nature of the immune response necessitates a comprehensive understanding of the spatiotemporal distribution of immune cells within the body, especially in tumors and tumor-draining lymph nodes (LNs) (*1,2*).

In recent years, molecular imaging techniques, such as positron emission tomography (PET), have emerged as powerful tools for enhanced understanding of the complex interactions between the immune system and cancer. Specifically, development of novel radiotracers targeting specific immune cells has introduced new means for non-invasive quantification of immune cell distribution within the tumor microenvironment (*3,4*). Among these radiotracers, ^18^F-arabinosyl guanine (^18^F-AraG) has garnered significant attention due to its selectivity towards activated T cells and its small molecular size.

^18^F-AraG is the ^18^F-labeled analog of the AraG, developed by Namavari et al (*5*) as a PET tracer for imaging T cell activation. The prodrug of AraG, Nelarabine, is FDA-approved for treatment of T cell acute lymphoblastic leukemia (T-ALL) and T cell lymphoblastic lymphoma and has been extensively studied for its selective T cell toxicity (*6,7*). Similar to AraG, ^18^F-AraG enters the cells via nucleoside transporters and gets phosphorylated by two enzymes: primarily by mitochondrial deoxyguanosine kinase (dGK), a critical enzyme for mitochondrial deoxyribonucleic acid (mtDNA) synthesis, and to a lesser extent by cytoplasmic deoxycytidine kinase (dCK). Tri-phosphorylated ^18^F-AraG can be incorporated into mtDNA or exit mitochondria and get dephosphorylated by sterile α motif and histidine-aspartate domain-containing protein 1 (SAMHD1), allowing ^18^F-AraG to be exported from the cell. Cells requiring increased DNA synthesis, such as activated T cells, exhibit downregulation of SAMHD1. The increased mitochondrial mass and mtDNA synthesis, and downregulation of SAMHD1 in activated T cells results in significantly higher uptake and retention of ^18^F-AraG by activated T cells compared to their unstimulated state, enabling use of ^18^F-AraG as an *in-vivo* probe to assess T cell activation (*8,9*).

^18^F-AraG’s uptake is not specific to activated T cells and ^18^F-AraG has considerable uptake in almost all organs, depending on expression and activity profiles of dGK and SAMHD1 in different cell types present in the organ (*9*). However, in the context of anti-tumor immune response evaluation, cancer cells other than malignancies of the T cell lineage are expected to have low uptake and retention of ^18^F-AraG, suggesting correlation of ^18^F-AraG tumor uptake with abundance and activation state of immune cells present in the tumor microenvironment. This is partly linked to the expression of SAMHD1 in cancer cells. Among a wide range of investigated cancer cell lines in Cancer Cell Line Encyclopedia, T-ALL cells show significantly lower expression of SAMHD1, which has been linked to AraG’s selectivity for activated T cells (*10*). Among human immune cells, T cells, macrophages, and dendritic cells have been shown to have the highest ^18^F-AraG uptake, with T cells showing the highest retention, in addition to significantly higher sensitivity to stimulation (*8*). Consistent with these findings, previous preclinical studies of ^18^F-AraG in a range of tumor models in mice have shown correlations of ^18^F-AraG tracer uptake with the frequency of CD8^+^ PD-1^+^ T cells in the tumors (*11*).

Previous studies utilizing ^18^F-AraG have demonstrated its potential in providing insights into the spatial distribution of tumor-infiltrating immune cells (*8*). However, the dynamic behavior of ^18^F-AraG within the body and its kinetic parameters remain unexplored. This study is the first report on dynamic imaging and kinetic modeling of ^18^F-AraG, as well as the first in-human report on using ^18^F-AraG in a non-small cell lung cancer (NSCLC) patient undergoing anti-PD-1 immunotherapy, aiming to enhance our understanding of its potential quantitative utility in predicting immunotherapy response, as well as its wider application in other disease conditions.

## MATERIALS AND METHODS

### Study Design

The study consisted of five subjects, including four healthy controls and one NSCLC patient. The demographics of the subjects are included in TABLE 1. The protocol was approved by the UC Davis Institutional Review Board (#1630355) and all participants provided written informed consent.

**TABLE 1.** Demographics of the study participants.

| ID | Group | Sex | Age range (y) | BMI (kg/m<sup>2</sup>) |
| --- | --- | --- | --- | --- |
| Sub01 | Control | F | 51–55 | 39 |
| Sub02 | Control | F | 71–75 | 27 |
| Sub03 | Control | F | 61–65 | 24 |
| Sub04 | Control | M | 41–45 | 30 |
| Sub05 | NSCLC | M | 51–55 | 25 |

The NSCLC patient had metastatic stage-IV NSCLC-adenocarcinoma of the left lower lobe (LLL) and right lower lobe (RLL), both diagnosed as primary tumors, histologically confirmed through bronchoscopy (PD-L1<1%). Both lesions were >1 cm in size and the LLL mass was sufficiently separated from other organs with known high ^18^F-AraG uptake, so that quantification was feasible. Quantification in the RLL was not feasible due to the proximity of the lesion to the liver. Fine needle aspiration biopsy from 1 of 3 mediastinal LNs was suspicious for metastatic carcinoma. The patient was enrolled in a clinical trial and was given pembrolizumab anti-PD-1 immunotherapy combined with pemetrexed-carboplatin chemotherapy every three weeks. Before the third immunotherapy session, the patient received a course of stereotactic body radiation therapy to the LLL lung with palliative intent. The oncologic history of the patient and the exclusion criteria of the study is provided in Supplementary Materials. The timeline of the medical interventions of the NSCLC patient with respect to the study is provided in SUPPLEMENTAL FIGURE 1.

### Radiotracer Formulation and Administration

^18^F-AraG was synthesized by Optimal Tracers (Sacramento, CA, USA). Participants received an intravenous (IV) bolus injection of a ∼189 MBq (mean: 189.0±12.6 MBq, range: 168.1‒200.6 MBq) dose of ^18^F-AraG. Estimated effective radiation dose was 2.9 ± 0.29 mSv and 3.1 ± 0.31 mSv for male and female subjects, respectively (*9*). No pre-medications were administered, and vital signs were recorded before and after imaging. All participants had either a follow-up visit or a call seven days after the scan to assess any adverse effects that could be attributed to the scan or administration of ^18^F-AraG.

### PET/CT Imaging

Subjects had 90-min total-body dynamic PET scans with ^18^F-AraG on the uEXPLORER scanner (United Imaging Healthcare, Shanghai, China) starting immediately prior to the IV bolus injection, except for Sub03 whose scan was terminated after 60 min due to patient discomfort and motion. A low-dose computed-tomography (CT) scan (dose-modulated, max 50 average mAs, 140 kVp, 10 mSv estimated effective radiation dose) was acquired prior to the PET scan of each subject for attenuation correction and anatomic correlation. The cancer patient was first scanned one day before the first immunotherapy session and again 14 days later after receiving the first cycle.

^18^F-AraG PET images were reconstructed using the vendor’s image reconstruction software, which used an iterative time-of-flight ordered-subset expectation maximization algorithm, with 4 iterations (20 subsets). Two sets of static reconstructions were performed on the 50‒60 min and the 60‒90 min frames of the dynamic scans, in addition to a 41-frame dynamic reconstruction using 15×2 s, 6×5 s, 3×20 s, 2×30 s, 3×60 s, 6×240 s, and 6×300 s frames, all using a 256×256 matrix with 2.344 mm isotropic voxels. The static reconstructions were used for visualization and localization of the volumes of interest (VOIs). The reconstructions used for data analysis were performed with no point spread function (PSF) modeling, whereas the reconstructions used for visualization included PSF modeling. Post-reconstruction kernel smoothing was applied to dynamic images, using a kernel matrix built from 4 consecutive composite frames of 10, 20, 30, and 30 min, including the 50 nearest neighbors of each voxel within a cubic 9×9×9 voxel space (*12*).

The initial staging of the cancer patient was based on a PET/CT scan performed 110 days before immunotherapy on a Discovery 690 scanner (General Electric Medical Systems, Milwaukee, WI, USA), acquired 56 min after a 375.7 MBq IV bolus injection of ^18^F-fluorodeoxyglucose (^18^F-FDG). Additionally, two sets of contrast-enhanced chest CT scans, with IV administration of 100 mL of Omnipaque 350, performed on a Revolution EVO scanner (General Electric Medical Systems, Milwaukee, WI, USA), two days before and 82 days after the first immunotherapy session, were used for tumor size assessment and initial therapy response evaluation.

### Image Analysis

Static PET/CT images were visualized in AMIDE. A semi-quantitative assessment was performed to identify the organs-of-interest with high uptake and ∼200 spherical VOIs were drawn on each dataset, over blood-pool (descending aorta and RV), cerebrum, choroid plexus, pituitary gland, salivary glands, thyroid, bone marrow, lungs, myocardium (RV and LV), spleen, liver, kidneys, femoral muscle, and LNs (axillary and pelvic). Additional VOIs were placed in regions of individual subjects where high focal uptake was observed. For the NSCLC patient, seven VOIs were placed on different sub-regions of the patient’s LLL tumor, to cover the tumor volume while including a variety of low-uptake and high-uptake sub-regions within the tumor. Moreover, four VOIs were placed on patient’s high-uptake mediastinal LNs, two of which were identified as enlarged, likely metastatic, LNs. For large organs (e.g., lungs, spleen, etc.), several VOIs were used to generate a segmented region. The image analysis was performed using an in-house developed code package in MATLAB R2021b. The standardized uptake value (SUV) was expressed as SUVmean in all organs, calculated from the average of the voxels within each segmented organ; except for LNs, which used SUVpeak due to their small size. In the tumor sub-regions, SUVmean was calculated directly from individual spherical VOIs placed on each sub-region. SUVpeak was defined as the mean of the eight hottest voxels within the VOI, equivalent of ∼100 mm^3^ volume.

### Kinetic Modeling

The descending aorta blood-pool was used as the input function for all organs, except for the lungs and the lung tumor sub-regions, for which the RV blood-pool was used. Three compartmental models were fitted on each time-activity curve (TAC) to select the model best describing the kinetics in each organ. This included the one-tissue compartmental model with four fitting microparameters (1T4P) of (*v_b_*, *K*_1_, *k*_2_, *t_d_*), two-tissue compartmental model with five fitting microparameters (2T5P) of (*v_b_*, *K*_1_, *k*_2_, *k*_3_, *t_d_*), and two-tissue compartmental model with six fitting microparameters (2T6P) of (*v_b_*, *K*1, *k*2, *k*3, *k*4, *t_d_*), in which *v_b_* is the fractional blood volume, *K*1, *k*2, *k*3, and *k*4 are the rate constants between model compartments, and *t_d_* is the time delay. Time delays were first jointly estimated with values ranging from −10 s to 50 s varied in steps of 1 s and then fitting was repeated for the other microparameters only, using the jointly-estimated time delays (*13*). The Levenberg-Marquardt algorithm was used for nonlinear least squares fitting, using nonuniform weighting factors defined based on the frame duration and decay factor (*14*). The Akaike information criterion (AIC) with a correction for small sample sizes was used to choose the model best fitting the data (*14*). The estimated time delays were used to delay-correct the blood-pool TACs for each organ and the datapoints from 14‒90 min were fitted with a biexponential function and extrapolated to 10 h post injection (p.i.) to model the tissue TACs at equilibrium time.

Tissue-to-blood ratio or SUVR was calculated from the ratio of the tissue activity concentration in each organ (*C_T_*) to the whole-blood activity concentration (*C_P_*). Net influx rate, 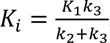, was calculated for the 2T model. Total volume of distribution, *V_T_*, was calculated as 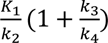 and 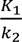 for the 2T6P and the 1T4P models, respectively (*15*). Since in the 2T6P model, SUVR at equilibrium time should reach the blood-volume-corrected *V_T_*, defined as *V_T_*(*v_b_*) = *v_b_* + *V_T_*(1 − *v_b_*); SUVR was estimated up to 10 h p.i., using the extrapolated blood-pool TACs, and compared to *V_T_*(*v_b_*) to evaluate the equilibrium time in different organs. Lastly, Logan plots (the linear regression between 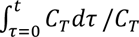 and 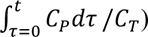) were generated and *K* was calculated from the slope of linear fits on data points after *K*^∗^ = 30 min (*16*).

To determine whether model parameters can be accurately estimated in presence of noise, practical identifiability analysis was performed, including calculation of normalized sensitivity curves, bias, standard deviation (SD), and root mean square error (RMSE) of the model parameters using 100 simulated noisy TACs (*17*). The practical identifiability analysis was repeated using the first 60-min datapoints only and bias, SD and RMSE of the model parameters were calculated in reference to the parameter estimations from the 90-min data fitting, to investigate kinetic modeling feasibility in future studies using 60-min dynamic scans. Detailed description of the kinetic modeling methods is provided in the Supplementary Materials and SUPPLEMENTAL FIGURE 2.

### Statistical Analysis

Spearman’s rank correlation coefficients were calculated in MATLAB R2021b between the *V_T_* and three parameters of SUV, SUVR, and *K*_*Logan*_ to investigate their use as surrogates of *V_T_*. Additionally, correlation coefficients of *V_T_*(*v_b_*) with SUVR and *K*_*Logan*_ were calculated for model validation and to investigate the effects from variabilities in tissue fractional blood volume. Hypothesis testing, comparing the pre-therapy and post-therapy scans of the NSCLC patient, was performed for the LLL tumor and the LNs in GraphPad Prism version 10.0, using a Wilcoxon paired signed rank test. P values <0.05 were set to determine statistical significance. No multiple comparison correction was included.

## RESULTS

The injections were well tolerated in all subjects, with no adverse effects and no clinically significant changes in vital signs.

### Biodistribution in Healthy Tissues

The SUV images (FIGURE 1) showed the highest uptake in liver, kidneys, and urinary bladder, followed by stomach wall, pancreas, salivary glands, thyroid, myocardium, spleen, bone marrow, choroid plexus, pituitary gland, ocular muscles, adrenal gland, and LNs. Prominent uptake was observed in pelvic and axillary LNs of all subjects. Cerebrum and cerebellum uptakes were negligible in all subjects, with SUVmean values below 0.06 and 0.08 for times >30 min p.i., respectively. The TACs from all healthy organs-of-interest investigated showed consistent trends (FIGURE 2 and SUPPLEMENTAL FIGURE 3), with a decrease in tissue SUVmean values for times approaching 90 min, except for liver and kidneys where radiotracer excretion effects are expected. Motion artifacts observed on TACs of two subjects towards the end of the scan time, particularly affected the quantification in the salivary glands of Sub01 and Sub03 and the pituitary gland in Sub03. Therefore, regions affected by motion were excluded from further analysis. Subjects showed variable levels of activity in their bowel; however, in most cases it was not possible to confirm whether the activity was in the lumen or the bowel wall, except for Sub03, in which cross sectional analysis suggested uptake in sections of the ascending colon wall (SUPPLEMENTAL FIGURE 4).

**FIGURE 1.**
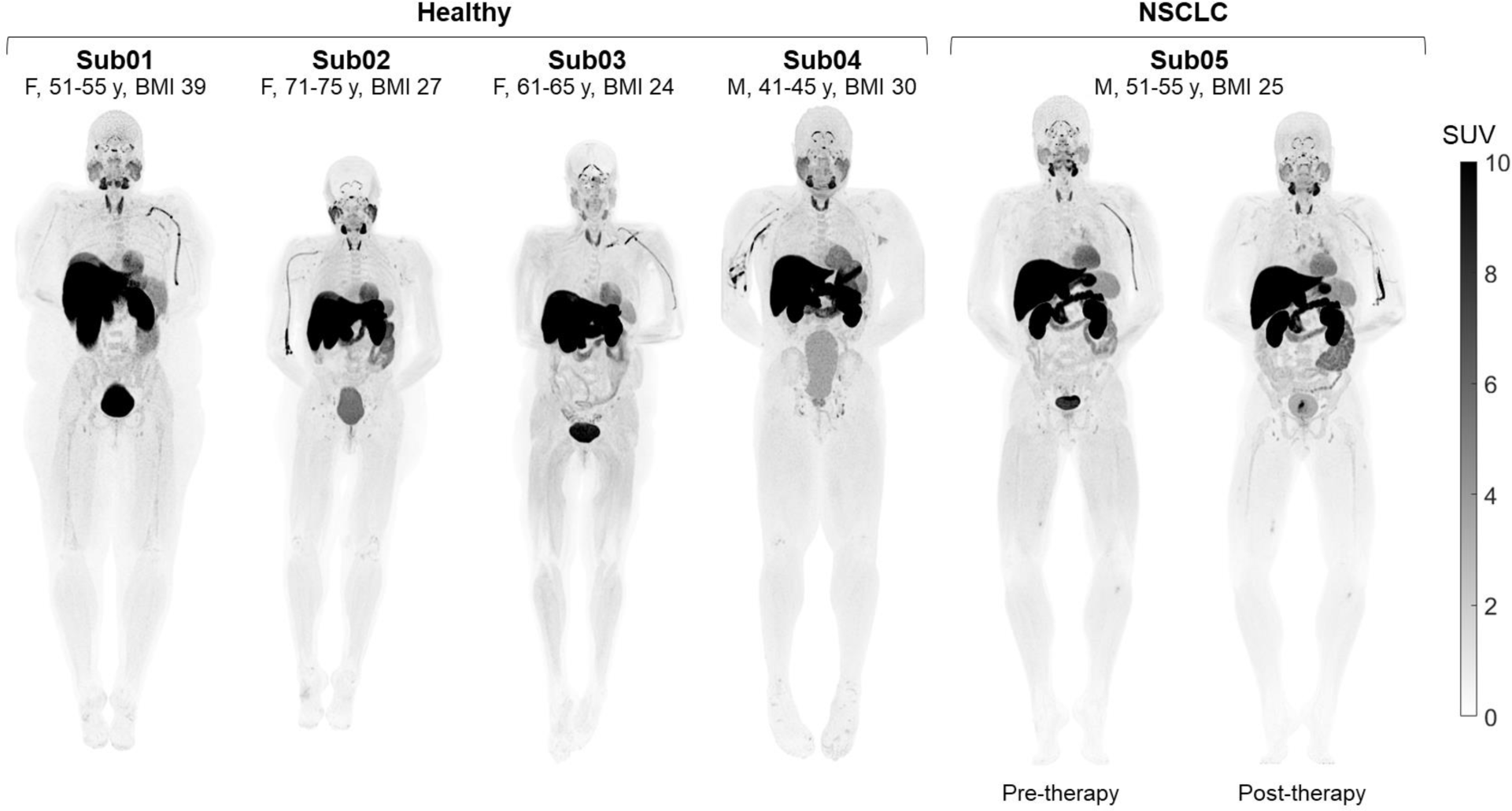
Maximum intensity projections of SUV images (50‒60 min p.i.) of the four healthy control subjects and one NSCLC patient scanned before and after immunotherapy.

**FIGURE 2.**
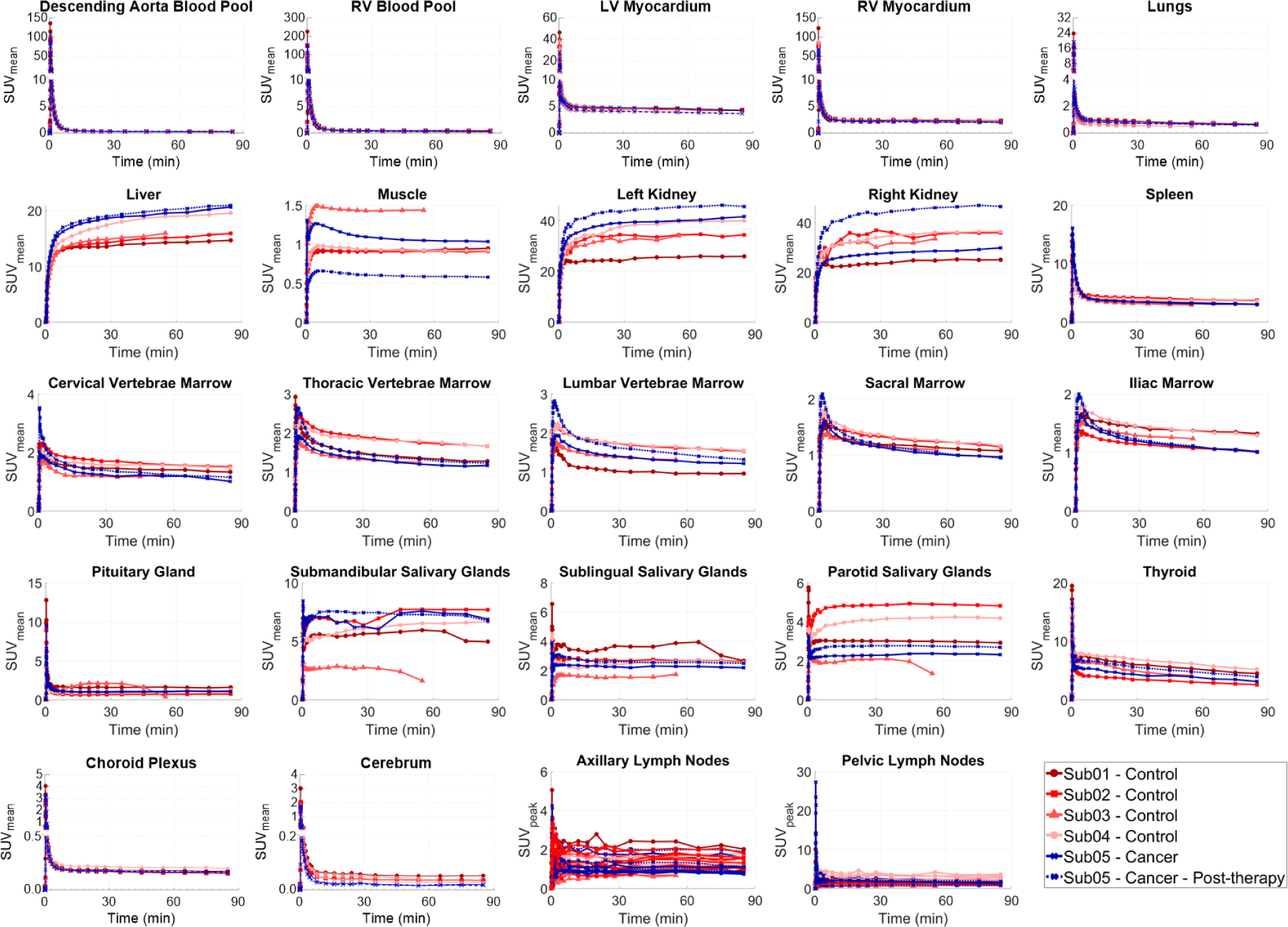
TACs of the healthy organs-of-interest analyzed in all subjects, showing the complete duration of the 90-min dynamic scans.

### Incidental Findings

Control subjects showed increased focal uptakes with altered kinetics at sites consistent with a range of pre-existing clinical conditions in their medical records. The bilateral underarm skin region showed an increased uptake and skin thickening compared to other skin regions in all subjects and the underarm skin uptake in Sub04, who had an asymptomatic chronic viral infection, was higher by a factor of 2 than other subjects (SUPPLEMENTAL FIGURE 5). Sub01 who had a history of pain in the arch of bilateral feet showed correspondingly high focal uptakes in the feet joints and Sub02 with history of osteoarthritis involving multiple joints showed focal uptakes in joints of hands and feet (SUPPLEMENTAL FIGURE 6). Sub03 with a complex middle cranial fossa base of skull meningioma showed correspondingly high uptake in the meningioma region (SUPPLEMENTAL FIGURE 7). The TACs of the tissues that showed increased uptake compared to their healthy state showed distinct kinetics, with plateauing or increasing uptake after 60 min p.i. (FIGURE 3 and SUPPLEMENTAL FIGURE 8).

**FIGURE 3.**
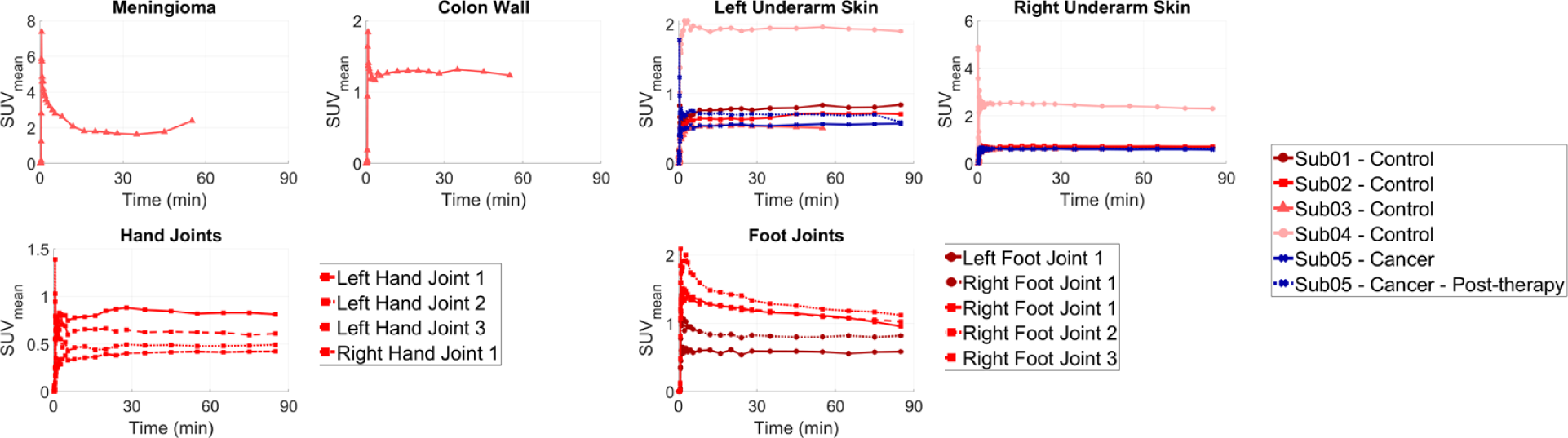
TACs of the organs with focally high uptake pattern in all subjects, showing the complete duration of the 90-min dynamic scans. The meningioma TAC was affected by head motion after 30 min p.i.

### Uptake in NSCLC Tumor and Mediastinal LNs

The LLL mass of the NSCLC patient showed a slight size reduction post-therapy, from 6.9×2.5 cm to 6.0×2.4 cm measured on the contrast-enhanced CTs (FIGURE 4). The two enlarged mediastinal LNs also showed slight size reduction post-therapy, from 2.1×1.4 cm to 1.5×0.9 cm and from 2.1×1.5 cm to 1.7×1.2 cm. Both the ^18^F-FDG pre-therapy scan and the ^18^F-AraG scans showed heterogeneous uptake in the LLL mass; however, the uptake patterns differed between the ^18^F-FDG and the ^18^F-AraG scans. The ^18^F-FDG scan showed substantially higher uptake in the posterior section of the tumor (SUVmax 14.8) compared to its other sub-regions (SUVmax 4.0), whereas the two ^18^F-AraG scans showed high-uptake sub-regions in both the anterior and the posterior sub-regions of the tumor with similar intensities and some sub-regions of the tumors with moderate ^18^F-FDG uptake showed no quantifiable uptake on the ^18^F-AraG scans (FIGURE 4). Furthermore, the two non-enlarged high-uptake mediastinal LNs on the ^18^F-AraG scans did not show quantifiable uptake on the ^18^F-FDG scan. The seven sub-regions of the LLL tumor selected for analysis (SUPPLEMENTAL FIGURE 9) and the four mediastinal LNs (SUPPLEMENTAL FIGURE 10), two of which were identified as enlarged LNs on the contrast-enhanced CTs showed similar trends of kinetics with a decreasing uptake between 30‒90 min (FIGURE 5 and SUPPLEMENTAL FIGURE 11). Comparing the SUVmean of the seven tumor sub-regions showed no trend in SUVmean changes post-therapy, varying from −12% to +45%. Similarly, the four mediastinal LNs showed a wide range of SUVpeak changes after therapy, varying from −22% to +112%.

**FIGURE 4.**
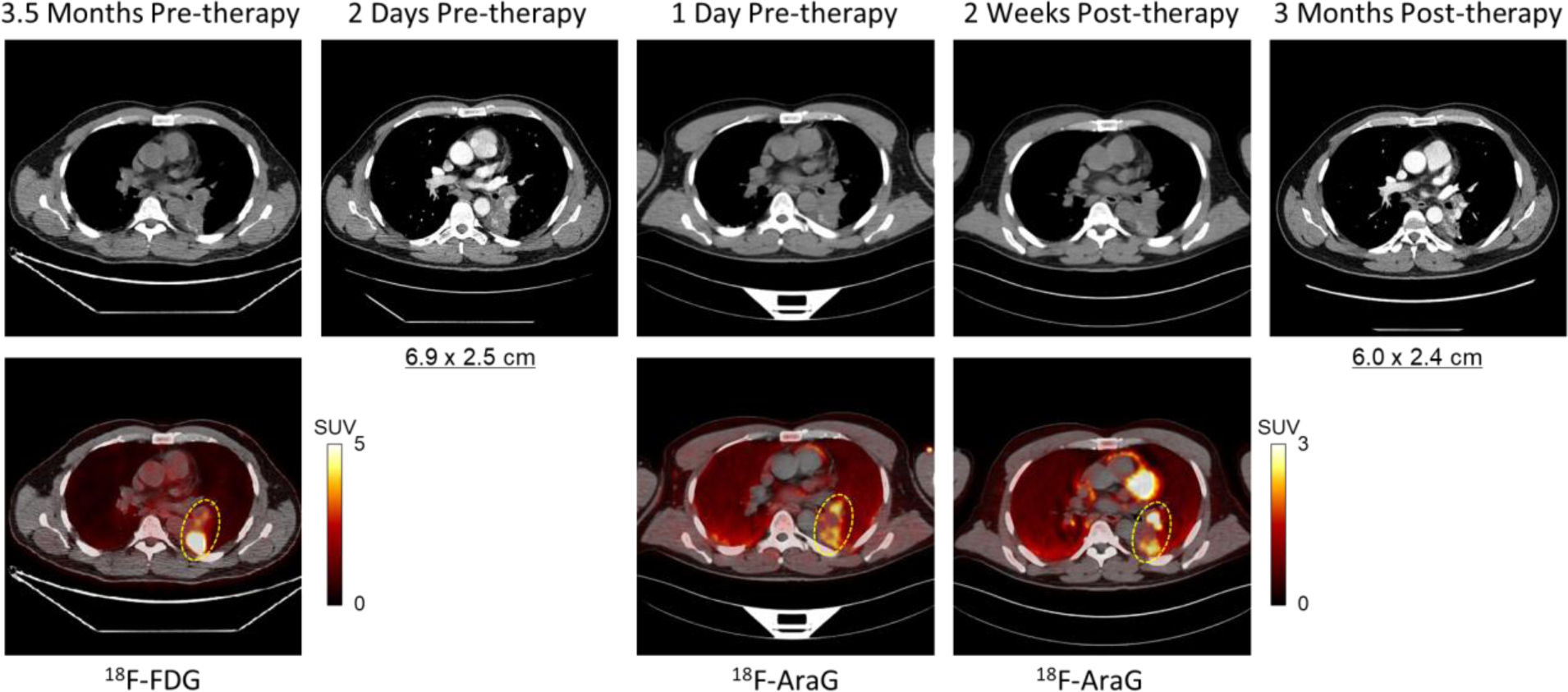
Transverse slices through the LLL mass of Sub05, comparing the initial ^18^F-FDG PET/CT at 3.5 months pre-therapy, contrast-enhanced CT at 2 days pre-therapy, ^18^F-AraG PET/CT at 1-day pre-therapy, ^18^F-AraG PET/CT at 2-weeks post-therapy, and the contrast-enhanced CT at 3-months post-therapy.

**FIGURE 5.**
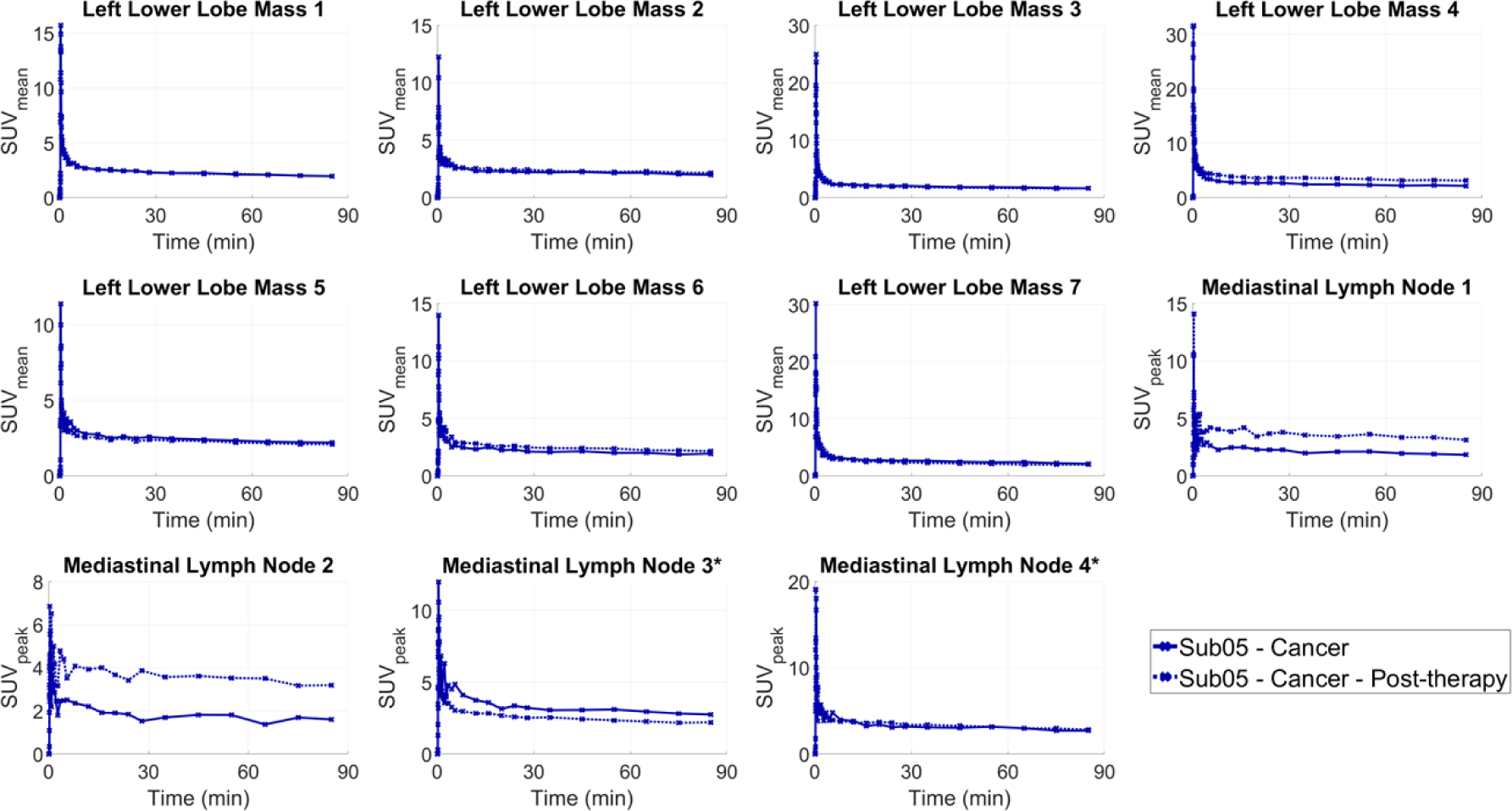
TACs of the seven sub-regions of the LLL mass and the four mediastinal LNs, showing the complete duration of the 90-min dynamic scans. The two enlarged LNs are marked with asterisks.

### Compartmental Modeling of the ^18^F-AraG Kinetics

PET kinetic modeling based on single input function one-tissue and two-tissue compartmental models with joint time-delay estimation successfully fitted the TACs in all analyzed organs-of-interest. In liver and RV myocardium, the initial peak of the TACs suffered from poor fitting. Furthermore, the LN TACs showed higher residual fitting errors due to the small number of voxels included in the analysis. AIC favored the 2T6P model over 2T5P and 1T4P models in all analyzed tumor sub-regions, mediastinal LNs, and the analyzed healthy organs-of-interest, except for axillary and pelvic LNs and RV myocardium (SUPPLEMENTAL FIGURE 12, SUPPLEMENTAL FIGURE 13, and SUPPLEMENTAL FIGURE 14). In 70 out of the 93 total analyzed axillary and pelvic LNs, the AIC favored the 2T6P model, whereas the 2T5P and the 1T4P were favored in 10 and 13 LNs, respectively (SUPPLEMENTAL FIGURE 15). Additionally, in the RV myocardium, AIC favored the 1T4P model in all subjects. The averaged parameter estimates for all analyzed organs-of-interest are summarized in SUPPLEMENTAL TABLE 1.

Normalized sensitivity plots (SUPPLEMENTAL FIGURE 16) showed that *K*_1_and *v_b_* reach their peak sensitivity during the first 1 min. While TAC sensitivity to *k*_4_showed a continuous increasing trend during the 90-min scans in all analyzed organs-of-interest, sensitivities to *k*_2_and *k*_3_varied in different organs (e.g. in LV myocardium, spleen, thyroid, and the LLL tumor, *k*_2_and *k*_3_ approached their maximum sensitivity during the first 15 min p.i., whereas, in RV myocardium, lungs, bone marrow, muscle, and joints, TAC sensitivities to *k*_2_and *k*_3_showed a continuous increasing trend during the 90-min scans).

Practical identifiability analysis with joint time-delay estimation showed low biases (−5.3%‒4.2%) in *V_T_* estimation in all analyzed organs-of-interest (TABLE 2). SD and RMSE of *V_T_* estimates were also low (2.4%‒10.3%) in most organs-of-interest (TABLE 3 and SUPPLEMENTAL TABLE 2), except for RV myocardium and muscle, where SD and RMSE of *V_T_* estimates increased up to 24.7% and 29.3%, respectively. The individual microparameter estimates showed higher variability compared to *V_T_*. Among all microparameters, *v_b_* showed the highest bias and variability. *K*_1_ showed low biases (−6.5%‒8.5%) in most organs, except in lungs and salivary glands, where *K*_1_ bias was 18.7% and –15.5%, respectively; however, SD and RMSE of *K*_1_ estimates were still higher than *V_T_* and were particularly high in the lungs and choroid plexus. In a number of organs, all rate-constant estimates showed low biases (within ±10%). This included LV myocardium, RV myocardium, spleen, thyroid, bone marrow, underarm skin, LLL tumor sub-regions, and mediastinal LNs; however, SD and RMSE of the rate constant estimates were only low (within ±10%) in RV myocardium.

**TABLE 2.** Estimated bias (%) of AIC-preferred model microparameters in analyzed organs-of-interest. The results show the averaged values among all subjects with 90-min dynamic scans. Results within <±10%, <±30%, and >±30% are marked with green, yellow, and red shadings, respectively.

| | $v_b$ | $K_1$ | $k_2$ | $k_3$ | $k_4$ | $V_T$ | $t_d$ |
| --- | --- | --- | --- | --- | --- | --- | --- |
| LV Myocardium | -0.5 | 1.6 | 4.4 | -1.5 | 0.5 | -0.5 | 0.8 |
| RV Myocardium | -0.3 | -0.1 | 0.2 | NA | NA | 4.2 | 0.0 |
| Lungs | -0.3 | 18.7 | 18.8 | -4.1 | 25.4 | -1.6 | -0.1 |
| Liver | 203.1 | 4.2 | 307.6 | 150.5 | 25.2 | 1.5 | 17.2 |
| Muscle | 12.3 | 8.5 | 159.1 | 153.0 | 4.3 | -5.3 | 28.1 |
| Spleen | 1.9 | -1.5 | -1.2 | 0.3 | -0.1 | -1.5 | -1.7 |
| Thyroid | 8.1 | -6.5 | -8.5 | 0.7 | 3.2 | 1.1 | 21.0 |
| Lumbar Vertebrae Marrow | 97.2 | 0.3 | 1.5 | -0.6 | -0.4 | -0.1 | 21.2 |
| Iliac Marrow | 108.3 | 1.0 | 6.7 | 1.8 | -0.8 | -3.1 | 16.4 |
| Parotid Salivary Glands | 114.8 | -15.5 | -27.0 | -7.6 | 22.1 | -0.8 | 49.4 |
| Choroid Plexus | 1.5 | -1.2 | -22.7 | 11.1 | 314.2 | 3.6 | -0.8 |
| Axillary and Pelvic LNs | 17.6 | -1.0 | 18.9 | 5.4 | 10.7 | 1.0 | -0.1 |
| Underarm Skin | 23.4 | -3.0 | 1.7 | 3.4 | 7.2 | -0.9 | -18.7 |
| Joints | -2.0 | 2.3 | 9.8 | 3.0 | -1.1 | -0.3 | -0.8 |
| LLL Tumor Sub-regions | -0.3 | -0.4 | 0.7 | 1.0 | -0.1 | 0.9 | 0.0 |
| Mediastinal LNs | 85.9 | 0.2 | 2.4 | 1.2 | 0.9 | 1.4 | -3.1 |

**TABLE 3.** Estimated standard deviation (%) of AIC-preferred model microparameters in analyzed organs-of-interest. The results show the averaged values among all subjects with 90-min dynamic scans. Results within <±10%, <±30%, and >±30% are marked with green, yellow, and red shadings, respectively.

| | $v_b$ | $K_1$ | $k_2$ | $k_3$ | $k_4$ | $V_T$ | $t_d$ |
| --- | --- | --- | --- | --- | --- | --- | --- |
| LV Myocardium | 7.3 | 15.8 | 38.8 | 17.5 | 11.1 | 4.0 | -15.2 |
| RV Myocardium | 3.3 | 7.9 | 6.4 | NA | NA | 11.6 | 0.0 |
| Lungs | 9.6 | 71.1 | 83.4 | 42.4 | 104.3 | 4.2 | 6.2 |
| Liver | 63.5 | 3.0 | 111.4 | 102.6 | 49.7 | 3.4 | 13.6 |
| Muscle | 48.0 | 23.6 | 245.6 | 334.7 | 69.1 | 24.7 | 50.6 |
| Spleen | 44.1 | 8.1 | 10.9 | 5.7 | 3.9 | 3.1 | 20.0 |
| Thyroid | 41.0 | 17.9 | 21.8 | 12.4 | 8.1 | 3.4 | 48.9 |
| Lumbar Vertebrae Marrow | 147.1 | 10.0 | 28.0 | 20.3 | 10.1 | 3.8 | 76.5 |
| Iliac Marrow | 163.6 | 11.7 | 45.8 | 31.9 | 12.9 | 4.3 | 43.9 |
| Parotid Salivary Glands | 176.2 | 22.5 | 33.9 | 16.8 | 30.2 | 3.6 | 94.9 |
| Choroid Plexus | 24.7 | 80.3 | 55.2 | 95.0 | 269.5 | 7.4 | 21.9 |
| Axillary and Pelvic LNs | 47.9 | 18.2 | 65.0 | 37.7 | 30.7 | 4.4 | 16.8 |
| Underarm Skin | 43.8 | 7.2 | 31.6 | 21.7 | 12.6 | 2.5 | -6.6 |
| Joints | 27.1 | 8.5 | 30.8 | 16.5 | 8.6 | 2.4 | 4.1 |
| LLL Tumor Sub-regions | 15.9 | 13.4 | 17.8 | 10.8 | 4.7 | 2.4 | 9.0 |
| Mediastinal LNs | 129.2 | 14.3 | 27.8 | 14.1 | 8.0 | 3.0 | -13.3 |

Repeating the practical identifiability analysis with setting the time-delays to their pre-estimated values substantially improved the bias and variability of the model microparameters in all organs-of-interest (SUPPLEMENTAL TABLE 3, SUPPLEMENTAL TABLE 4, and SUPPLEMENTAL TABLE 5). Furthermore, repeating the practical identifiability analysis with joint time-delay estimation and only using the datapoints from the first 60 min for model fitting showed very similar bias, SD, and RMSE in all parameter estimates compared to using the whole 90-min datasets (SUPPLEMENTAL TABLE 6, SUPPLEMENTAL TABLE 7, and SUPPLEMENTAL TABLE 8).

### Tissue-to-Blood Ratio and Logan Plot as Surrogates for *V_T_*

SUVR curves showed a plateauing trend towards the end of the 90-min dynamic scans in most analyzed organs-of-interest (SUPPLEMENTAL FIGURE 17, SUPPLEMENTAL FIGURE 18, and SUPPLEMENTAL FIGURE 19), except for liver and kidneys, which showed an increasing trend, and thyroid which showed a decreasing trend. The SUVRs showed increases (+4% to +71%) in all seven sub-regions of the LLL tumor post-therapy during 60‒90 min p.i. The two enlarged mediastinal LNs showed decreased SUVRs post-therapy (−33% and −10%), while the two non-enlarged mediastinal LNs showed an increasing trend post-therapy (48% and 82%). Comparing the 10-h extrapolated modeled SUVR curves to the *V_T_*(*v_b_*) showed smaller differences between SUVR curves and *V_T_*(*v_b_*) at 10 h p.i. compared to 90 min p.i. in all investigated organs-of-interest (SUPPLEMENTAL FIGURE 20 and SUPPLEMENTAL FIGURE 21). Furthermore, Logan plots showed a linear slope in all analyzed organs-of-interest after 30-min p.i. (SUPPLEMENTAL FIGURE 22, SUPPLEMENTAL FIGURE 23, and SUPPLEMENTAL FIGURE 24).

Both *K*_*Logan*_ and SUVR values from 60‒90 min p.i. were highly correlated with *V_T_*(*v_b_*) in all organs-of-interest, with correlation coefficients within 0.80‒1.00 and 0.85‒1.00, respectively (FIGURE 6). Similarly, *K*_*Logan*_ and SUVR values were highly correlated with *V_T_* in most organs-of-interest, showing slightly lower correlation coefficients (<5%) compared to *V_T_*(*v_b_*) correlations, except for LV and RV myocardium and parotid salivary glands, in which correlation coefficients with *V_T_* were substantially lower than with *V_T_*(*v_b_*). SUVs from 60‒90 min p.i. also showed moderate to strong correlations with *V_T_* in some organs, including thyroid, bone marrow, LNs, underarm skin, and joints, with correlation coefficients within 0.74‒1.00. However, in other organs, correlations were weak and in case of lungs, the SUVmean was negatively correlated with *V_T_*. Correlations of *K*_*i*_ values obtained from the 2T6P and 2T5P models with SUVR also showed inconsistent results in the analyzed organs-of-interest, ranging from negative correlations in some organs (e.g. myocardium, lungs, and thyroid) to moderate or strong positive correlations in others (e.g. joints, LNs, and tumor sub-regions).

**FIGURE 6.**
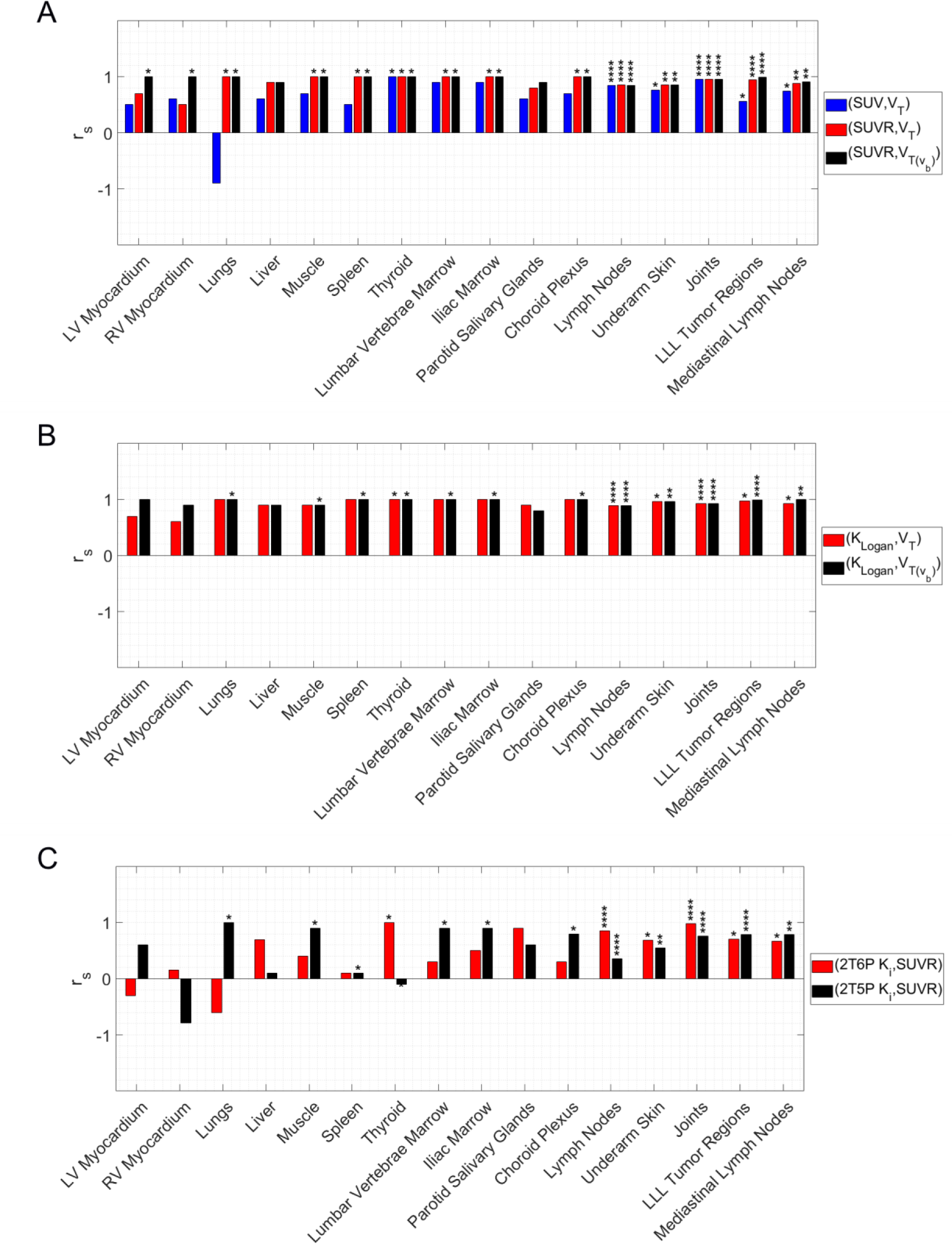
Spearman’s rank correlation coefficients (***r_s_***) calculated (A) between (SUV, ***V_T_***), (SUVR, ***V_T_***), and (SUVR, ***V_T_*(*v_b_***)), (B) between (***K_Logan_*, *V_T_***) and (***K_Logan_*, *V_T_v_b_***)), and (C) between (2T6P ***K_i_***, SUVR) and (2T5P ***K_i_***, SUVR) in different organs-of-interest. Asterisks shown above individual bars show the significance level of the Spearman’s rank p-values (*p<0.05, **p<0.01, ***p<0.001, ****p<0.0001).

Lastly, comparing the pre- and post-therapy scans of the NSCLC patient, SUVR, *K*_*Logan*_, and *V_T_* showed significant (p=0.016) increases in all seven analyzed sub-regions of the LLL tumor after therapy, in addition to significantly (p= 0.047) increased *k*_3_ and significantly (p=0.016) decreased *k*_4_(FIGURE 7). Changes of *K*_1_and *k*_2_were variable in different sub-regions of the tumor. The changes of SUVR, *K*_*Logan*_, and *V_T_* of the mediastinal LNs were different between the two enlarged LNs and the two non-enlarged LNs, with decreasing and increasing trends post-therapy, respectively (SUPPLEMENTAL FIGURE 25). Lastly, the analyzed axillary and pelvic LNs of the NSCLC patient also showed a significant (p<0.0001) increasing trend in *k*_3_ post-therapy, as well as a significant (p=0.045) decreasing trend in SUVR; however, no significant changes in *K*_*Logan*_, and *V_T_* were observed (SUPPLEMENTAL FIGURE 26).

**FIGURE 7.**
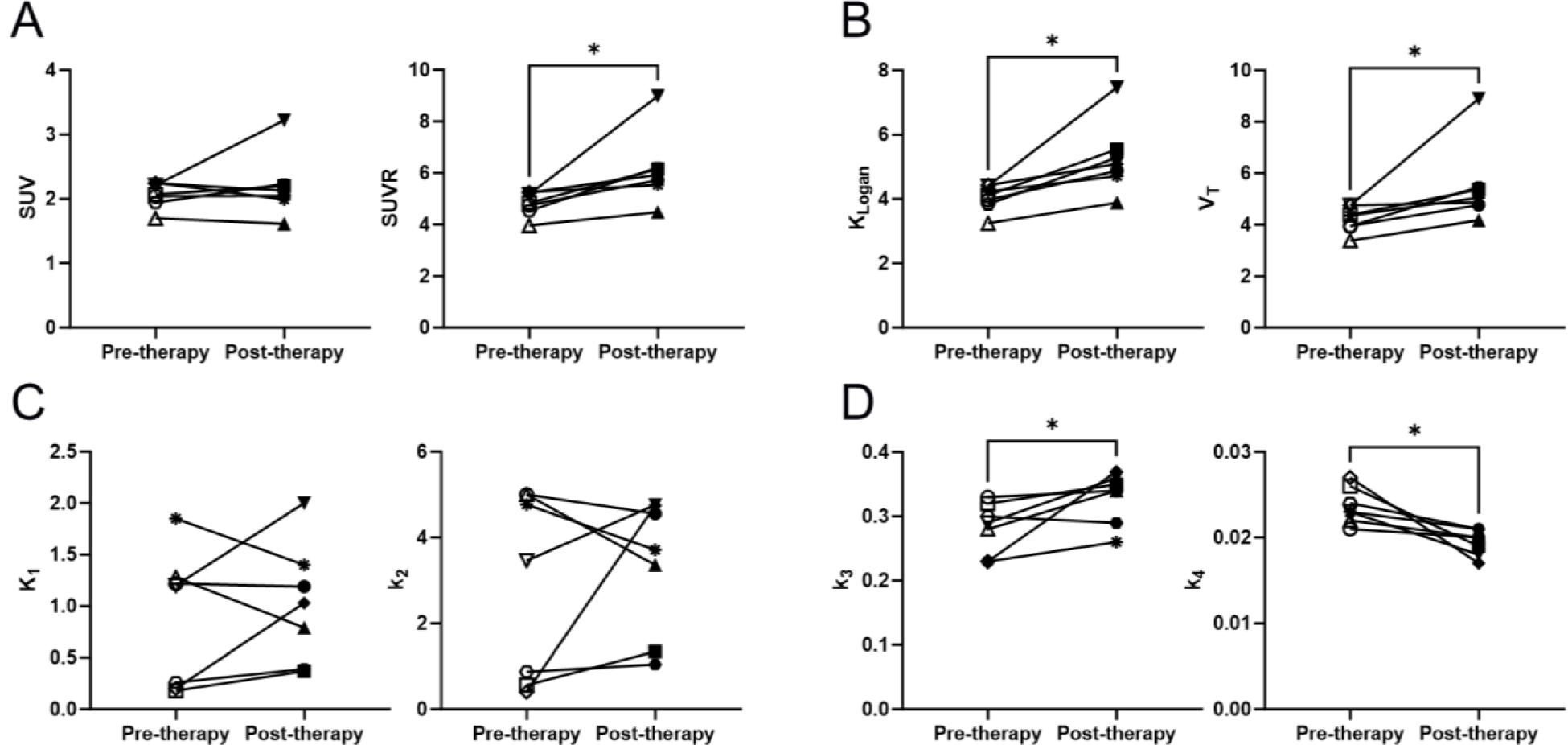
Changes in seven sub-regions of the LLL tumor post-therapy, using (A) SUVmean or SUVR values without kinetic modeling, (B) ***K_Logan_*** from Logan plots or ***V_T_*** from 2T6P compartmental modeling as macroparameters with high identifiability, (C) 2T6P rate constants between blood and first compartment (***K*_1_** and ***K_2_***), and (D) 2T6P rate constants between first and second compartments (***k_3_*** and ***k_4_***).

## DISCUSSION

Total-body late-timepoint biodistributions were in good agreement with previous static ^18^F-AraG scans in healthy volunteers (*9*). However, the high sensitivity of total-body PET resulted in visualization of a notably larger number of LNs in all subjects of this study, despite the ∼40% reduction in injected radiation dose compared to previous in-human studies. This is of particular interest for evaluating the immune response to therapeutic interventions in cancer patients, due to the critical role of tumor-draining LNs, but also in other disease conditions that stimulate the immune system.

Tumor ‘immune contexture’ has been previously proposed to describe the nature, density, immune functional orientation and distribution of immune cells within the tumor and has been associated with long-term survival and treatment response prediction (*18,19*). The tumor immune contexture has been increasingly used for classification of tumors into major categories of hot, altered, and cold tumors (*18*). In the context of using ^18^F-AraG imaging as a biomarker for immunotherapy response evaluation, three major factors are considered to play key roles in this study: first, the initial ^18^F-AraG scan serving as a biomarker for quantification of the baseline antitumor immune response and tumor immune contexture classification; second, changes in tumor uptake in follow-up post-therapy ^18^F-AraG scans serving as biomarkers of therapy-induced alterations in tumor microenvironment; and third, quantification in tumor-draining LNs as important sites of antigen presentation and T cell activation. While ^18^F-AraG SUV images could serve for baseline tumor classification as previously shown in preclinical studies (*11*), parameters obtained by kinetic modeling, or their surrogates, are expected to provide higher accuracy in quantification of the uptake especially when different scans are compared, primarily due to differences in time-varying blood input function and its subsequent effects on tissue SUVs. Analyzing seven sub-regions of a NSCLC tumor in this study has shown promising results in evaluating the therapy-induced changes in tumor microenvironment. Despite the variability observed in tumor SUVmean changes post-therapy, the SUVR, *K*_*Logan*_, and *V_T_* showed increasing trends in all analyzed sub-regions of the tumor. Furthermore, the significant increasing trend in *k*_3_and the decreasing trend in *k*_4_observed in post-therapy scans of the tumor are in good agreement with the expected immunotherapy-induced increase in T cell activation, leading to higher dGK and lower SAMHD-1 expression in the activated T cells infiltrating the tumor (*8,9*). These findings are also consistent with the slight tumor size reduction evidenced on CT scans, suggesting initial effectiveness of therapy in the tumor.

The differences observed in tumor uptake patterns between ^18^F-FDG and ^18^F-AraG are consistent with previous preclinical studies (*8*) and can be attributed to fundamental differences in the source of uptake in ^18^F-FDG and ^18^F-AraG scans. The large contribution of tumor cell metabolism to the ^18^F-FDG signal prevents accurate quantification of the immune contexture within the tumor microenvironment, whereas a major fraction of ^18^F-AraG uptake in NSCLC tumors is expected to originate from the immune cells in the tumor (*8*). However, the 3.5 months interval between the ^18^F-FDG and the ^18^F-AraG scans in this study may have also contributed to the progression of the tumor and its metastatic LNs.

The substantial increase of SUVR, *K*_*Logan*_, and *V_T_* observed in the two non-enlarged mediastinal LNs and the decreasing trend observed in the two enlarged mediastinal LNs may be explained by differences in therapy response in uninvolved and involved regional LNs (*20*); however, it requires further investigation in a larger cohort of patients, especially considering the slight size reduction of the two enlarged nodes on post-therapy CT images. Given that the practical identifiability analysis did not show significantly increased bias or variability in the mediastinal LNs compared to other organs, the current findings suggest that quantitative response assessment in tumor-draining LNs might require multiple longitudinal scans of the patient post-therapy to consider effects from T cell trafficking (*2*). Interestingly, the axillary and pelvic LNs of the patient showed a significant increasing trend in *k*_3_, which is consistent with the expected global effect of anti-PD-1 immunotherapy on peripheral T cells, distant from the primary tumor site.

Promising kinetic modeling results obtained in all investigated organs-of-interest suggest that ^18^F-AraG imaging can be used as a quantitative biomarker for a wide range of applications, such as graft-versus-host disease, arthritis, multiple sclerosis, and viral infections. This is further supported by the incidental findings of the study, showing focal uptakes in potential sites of immune infiltration, such as osteoarthritis joints, colon wall, meningioma, and the underarm skin. The AIC analysis favored the reversible two-tissue compartmental model compared to other models across various organs-of-interest with the tested high-temporal-resolution framing protocol. The kinetic modeling was validated through the strong correlations observed between (SUVR, *V_T_*(*v_b_*)) and (*K*_*Logan*_, *V_T_*(*v_b_*)) in all organs-of-interest. Furthermore, proximity of *V_T_*(*v_b_*) estimates to extrapolated SUVR curves at 10 h p.i. is suggestive of *V_T_*(*v_b_*) estimation accuracy and the remaining differences observed between the *V_T_*(*v_b_*) and the converged SUVR curves at equilibrium are possibly from biexponential fitting errors on whole-blood TACs, which requires future validation using delayed imaging. Comparing the extrapolated SUVR curves in different organs suggests that equilibrium may be reached after 4-h p.i. in the analyzed organs-of-interest with substantial differences in the organ equilibrium times, as expected.

The correlations between *K*_*Logan*_ and SUVR values with *V_T_* were generally strong, suggesting that these measures could serve as surrogates for *V_T_* estimation without kinetic modeling. However, their use may require careful attention in organs with high blood-volume fraction, such as myocardium. SUVmeans from 60-90 min p.i. showed only moderate to strong correlations with *V_T_* in a few organs, including the tumor sub-regions, which indicates their potentially limited utility for quantification of tracer uptake. The present study recommends use of SUVR over SUV in static scans, where kinetic modeling or graphical analysis methods may not be feasible. The negative correlation observed between lung SUVmean and *V_T_* requires further investigation with a larger sample size, but also highlights the potential misleading nature of SUVs in certain cases.

The differences observed between the two *K*_*i*_ estimates and the variability of the correlation results in different organs suggest that *k*_4_ effects may not be negligible in many organs-of-interest. In tissues where activated T cells are expected to be dominating factors in the tracer kinetics, such as the tumor, LNs, or the investigated joints of the two control subjects in this study, the difference between the two *K*_*i*_ estimates becomes smaller and stronger correlations with SUVR are observed. However, even in such cases, SUVR values still correlate better with *V_T_* and *V_T_*(*v_b_*) and using a reversible model is recommended.

The practical identifiability analysis suggests that *V_T_* can be estimated reliably in all organs-of-interest with low bias and variability. The individual rate-constant estimates also showed low biases, but had larger variabilities than *V_T_* particularly in healthy organs, which may require larger sample sizes when used for hypothesis testing. The improved results obtained by fixing the time-delay values suggest that variabilities in time-delay estimation may largely affect the input function properties, leading to increased variabilities in microparameter estimation. Therefore, future studies should investigate use of improved time-delay estimation methods. While a continuous increasing trend in *k*_4_ sensitivity was observed during the 90-min scans in all organs-of-interest, the identifiability analysis results obtained with 60-min TACs suggest sufficient sensitivity for *k*_4_in the first 60 min for reducing the dynamic scan duration in future studies.

This pilot study had some limitations. The small sample size and the inclusion of a single NSCLC patient warrant further investigations in larger cohorts. Future studies should also investigate the effect of tracer uptake by immune cells present in whole-blood on the kinetic modeling by longitudinal blood sample collection, as well as quantifying the tracer metabolites in blood and their effects on kinetic parameters. Lastly, the currently used single-input-function compartmental modeling may not be optimal for a number of organs, such as liver, RV myocardium, and certain types of lung tumors, and the models can be further optimized by incorporating additional factors, such as dual-input-function, dispersion, and air fraction correction.

## CONCLUSION

In conclusion, this study demonstrates the potential of ^18^F-AraG dynamic imaging as a non-invasive biomarker for evaluating the immune response to therapeutic interventions in cancer patients and underscores the importance of quantification methods, such as kinetic modeling or graphical methods, for accurate assessment of immune contexture and therapeutic response prediction. Increasing trends were observed in SUVR, *K*_*Logan*_, and *V_T_* in all analyzed sub-regions of the tumor post-therapy, whereas SUVmean changes varied in different sub-regions of the tumor. The correlations between *K*_*Logan*_ and SUVR values with *V_T_* show promise as surrogates for *V_T_* estimation, especially in organs with low blood volume fraction. In addition, the study suggests that the dynamic ^18^F-AraG PET scans could potentially be shortened to 60 minutes, while maintaining quantification accuracy for all organs-of-interest. Lastly, the incidental findings of the study which were consistent with the medical history of the subjects, further highlight potentially wider applications of the tracer in investigating the role of T cells in the immunopathogenesis of diseases.

## FINANCIAL DISCLOSURE

This work was supported by CellSight Technologies and National Institutes of Health grants R01CA206187 and R35CA197608.

## DISCLAIMER

Jelena Levi is employed by CellSight Technologies. CellSight Technologies Inc. is commercializing ^18^F-AraG. No other potential conflict of interest relevant to this article was reported.

## Data Availability

All data produced in the present study are available upon reasonable request to the authors.

## ACKNOWLEDGMENTS

The authors would like to thank Dr. Benjamin Spencer for technical and scientific support during this project. Furthermore, the authors thank Lynda Painting (research coordinator) and the team of technologists at EXPLORER Molecular Imaging Center for their assistance in the study.

## Notes

### Competing Interest Statement

Jelena Levi is employed by CellSight Technologies. CellSight Technologies Inc. is commercializing 18F-AraG. No other potential conflict of interest relevant to this article was reported.

### Author Declarations

Ethics committee and the Institutional Review Board of the University of California Davis (Davis, CA, USA) gave ethical approval for this work. The IRB number is #1630355.

### Summary of Updates

Results section on incidental findings was updated.

