## Supplementary material for "Total-body Dynamic Imaging and Kinetic Modeling of ^18^F-AraG in Healthy Individuals and a Non-Small Cell Lung Cancer Patient Undergoing Anti-PD-1 Immunotherapy": Supplmental Materials

### Supplementary Materials

#### Study's Exclusion Criteria

The exclusion criteria for all participants were subjects with serious comorbidities, history of recent COVID-19 infection within the last 2 months or history of COVID-19 requiring hospitalization with lung injury, subjects with a diagnosis of immunodeficiency or receiving systemic steroid therapy or any other form of immunosuppressive therapy within 7 days prior to the scan, or subjects receiving therapy with nucleoside analogs. The NSCLC patient had no history of prior treatment with anti-PD-1/PD-L1 immunotherapy.

#### NSLC Patient's Oncologic History

Patient was admitted to the emergency room presenting large volume (~200 mL) hemoptysis. Patient was evaluated and was stable to be discharged home with doxycycline for pneumonitis. After a recurrent episode of hemoptysis on the same evening, the patient was admitted to the intensive care unit for close monitoring. Chest CT scan was performed indicating a 4.9 × 3.7 cm LLL mass with left hilar adenopathy, along with interstitial lung disease of the right lower lobe. After two days, the patient was sedated and intubated before bronchoscopy with EBUS and fine needle aspiration of LLL mass and LNs, but the bronchoscopy results were non-diagnostic. Complicated by significant LLL bleeding during bronchoscopy, the patient was transferred immediately to interventional radiology for left bronchial artery embolization. The patient underwent brushing of the LLL with bronchoalveolar lavage, which also did not return any malignant cells. The patient was discharged after two days.

Ten days later, patient underwent an $^{18}$F-FDG PET/CT scan, which showed bilateral pulmonary nodules with several FDG-avid mediastinal and left hilar lymph nodes. Patient underwent a second bronchoscopy. Ion fine needle aspiration of the LLL mass showed adenocarcinoma, PD-L1<1%, FoundationOne revealing KRAS G12D and SMAD4 A118V. Three lymph nodes from stations 4L, 7, and 11L were also biopsied through fine needle aspiration, one which (station 4L) was suspicious for metastatic carcinoma. Patient's care was discussed with the Thoracic Multidisciplinary Tumor Board. The recommended plan was to pursue RLL biopsy and if the RLL biopsy morphologically matches the LLL tumor histopathology and/or the molecular phenotype is concordant with the LLL NGS (e.g., KRAS G12D presence), then the patient is likely to harbor metastatic stage-IV NSCLC. RLL biopsy was performed and revealed non-small cell carcinoma with glandular features. Clinical trial candidacy was discussed with the patient for a randomized phase

II/III trial of modern immunotherapy based systemic therapy with or without stereotactic body radiation therapy for PD-L1-negative, advanced non-small cell lung cancer.

**Kinetic Modeling Methods**

In this study, the kinetics of $^{18}$F-AraG is generally described by a reversible two-tissue (2T) compartmental model (SUPPLEMENTAL FIGURE 2), consisting of five microparameters of $(K_1, k_2, k_3, k_4, v_b)$. Time delay $(t_d)$ is included as the sixth microparameter and is estimated jointly with other model microparameters. $C_T(t)$ is the activity concentration of the tissue-of-interest measured on dynamic PET images as a function of time, $t$. $C_P(t)$ is the activity concentration of $^{18}$F-AraG in arterial blood plasma and the input function (IF) to the system. The subscript $P$ was used to represent the plasma activity concentration conventionally used as model input function; whereas an image-derived input function was used in this study (the RV blood-pool for the lungs and the lung tumor sub-regions and the descending aorta blood-pool for all the other organs). $C_1(t)$ and $C_2(t)$ are the two tissue compartments, representing concentrations of free $^{18}$F-AraG and tri-phosphorylated $^{18}$F-AraG ($^{18}$F-AraG-TP) in tissue cells, respectively. $K_1$ ($mL/min/mL_{Tissue}$) and $k_2$ ($min^{-1}$) are the rates of $^{18}$F-AraG transport between arterial blood plasma and tissue cells and are blood-flow de. $k_3$ ($min^{-1}$) and $k_4$ ($min^{-1}$) are $^{18}$F-AraG phosphorylation and dephosphorylation rates, respectively and $v_b$ is the fractional blood volume in tissue.

The tissue activity concentration measured by PET is described by: $C_T(t) = v_b C_{WB}(t) + (1 - v_b)\big(C_1(t) + C_2(t)\big)$, where $C_{WB}$ is the activity concentration of the tracer in whole blood, including any uptake by blood cells. The blood plasma fraction $(C_P/C_{WB})$ is assumed to be one in this study, for simplification. This 2T compartment system can be described by two first-order linear differential equations:

$$\frac{dC_1(t)}{dt} = K_1 C_P(t) - (k_2 + k_3)C_1(t) + k_4 C_2(t),$$

$$\frac{dC_2(t)}{dt} = k_3 C_1(t) - k_4 C_2(t)$$

and the solution to such 2T model is:

$$C_1(t) = \frac{K_1}{(\alpha_2 - \alpha_1)}[(k_4 - \alpha_1)e^{-\alpha_1 t} + (\alpha_2 - k_4)e^{-\alpha_2 t}] * C_p(t),$$

$$C_2(t) = \frac{K_1 k_3}{(\alpha_2 - \alpha_1)}(e^{-\alpha_1 t} + e^{-\alpha_2 t}) * C_p(t),$$

in which

$$\alpha_1 = \frac{(k_2+k_3+k_4)-\sqrt{(k_2+k_3+k_4)^2-4k_2k_4}}{2},$$

$$\alpha_2 = \frac{(k_2+k_3+k_4)+\sqrt{(k_2+k_3+k_4)^2-4k_2k_4}}{2}.$$

This is a special case of the general solution of an $n$-tissue compartmental model, in which the total concentration of tracer in tissue compartments can be described by the sum of convolution integrals as

$$\sum_{i=1}^{n} C_i(t) = IRF(t) * C_P(t) = \sum_{i=1}^{n} \phi_i e^{-\theta_i t} * C_P(t),$$

in which $IRF(t)$ is the impulse response function of the system described by sum of exponential functions. This demonstrates the susceptibility of PET static imaging to inter-scan variabilities in the blood IF, as tissue activity concentrations measured in PET not only have direct a contribution from blood IF due to tissue blood volume fraction, but also the total concentration of tracer in tissue compartments is the result of a convolution of the tissue's IRF by the time-variant blood IF.

Total volume of distribution ($V_T$) is a macroparameter often used in reversible compartmental models, which is the step response function (SRF) of the system at $t \to \infty$, when the system is at equilibrium and no further changes in the compartments are expected ($\lim_{t\to\infty} \frac{dC_1(t)}{dt} = 0$ and $\lim_{t\to\infty} \frac{dC_2(t)}{dt} = 0$). $V_T$ in a reversible 2T model can be derived from

$$V_T = \frac{K_1}{k_2}(1 + \frac{k_3}{k_4})$$

and is the sum of the volume of distributions in each compartment and the converged value of $(C_1 + C_2)/C_P$ at equilibrium time ($t \to \infty$). The volume of distribution in the two tissue compartments are:

$$V_1 = \lim_{t\to\infty} \frac{C_1(t)}{C_P(t)},$$

$$V_2 = \lim_{t\to\infty} \frac{C_2(t)}{C_P(t)}.$$

By setting $\frac{dC_1(t)}{dt}$ and $\frac{dC_1(t)}{dt}$ to zero at equilibrium time, the volumes of distribution in each compartment can be calculated as:

$$V_1 = \frac{K_1}{k_2},$$

$$V_2 = \frac{K_1 k_3}{k_2 k_4}.$$

Tissue-to-blood ratio (TBR) or SUVR (measured even before equilibrium) is often proposed as a surrogate for $V_T$, when kinetic modeling is not possible in static scans. This can particularly hold true in tissues with low fractional blood volume ($v_b \ll 1$) since $C_T(t) \approx C_1(t) + C_2(t)$ and $\lim_{t\to\infty} C_T(t)/C_P(t) \approx V_T$. However, in tissues with high fractional blood volume (e.g., lungs or myocardium), SUVR can be expected to have high correlations only with the blood-volume corrected $V_T$, defined as

$$V_{T(v_b)} = v_b + (1 - v_b)V_T.$$

In this study, to validate the kinetic model in different tissue types, the activity concentrations in each tissue compartment ($C_1$ and $C_2$) were estimated (based on the estimated model microparameters and the extrapolated IF up to 10 h p.i.) and corrected for fractional blood volume by multiplying them with $(1 - v_b)$ and their ratio to blood activity concentration were compared to blood-volume corrected volumes of distribution in each compartment corrected ($V_{ND(v_b)} = v_b + \frac{K_1}{k_2}(1 - v_b)$ in the first compartment and $V_{S(v_b)} = v_b + \frac{K_1 k_3}{k_2 k_4}(1 - v_b)$ in the second compartment). Subsequently, the blood-volume corrected total volume-of-distribution ($V_{T(v_b)}$) was compared to the modeled SUVR at 10 h p.i. (SUPPLEMENTAL FIGURE 20 and SUPPLEMENTAL FIGURE 21).

Lastly, the reversible 2T model may not be suitable for describing the $^{18}$F-AraG kinetics in all tissue types. As an example, dual blood supply of liver requires adjustments to the compartmental model to incorporate two IFs. Furthermore, RV myocardium PET signal may contain spill-over from the RV blood-pool, which has not been accounted for in the currently use compartmental modeling.

### Supplementary Tables

**SUPPLEMENTAL TABLE 1.** Micro- and macro-parameter estimates of the AIC-preferred model microparameters in analyzed organs-of-interest, calculated from model fitting on 90-min data. The results show the average ± standard deviation of values among all subjects with 90-min dynamic scans.

| | $v_b$ | $K_1$ | $k_2$ | $k_3$ | $k_4$ | $V_T$ | $t_d$ |
|---|---|---|---|---|---|---|---|
| LV Myocardium | 0.30±0.07 | 0.66±0.56 | 1.40±2.03 | 0.45±0.22 | 0.020±0.004 | 16.5±1.9 | -0.6±0.5 |
| RV Myocardium | 0.71±0.09 | 0.22±0.06 | 0.01±0.00 | NA | NA | 22.1±9.6 | -7.0±0.7 |
| Lungs | 0.15±0.02 | 0.17±0.11 | 2.89±1.94 | 0.61±0.15 | 0.025±0.002 | 1.5±0.1 | 3.2±0.4 |
| Liver | 0.00±0.00 | 0.48±0.07 | 0.11±0.20 | 1.13±1.02 | 0.418±0.531 | 75.9±18.3 | 10.6±1.8 |
| Muscle | 0.01±0.01 | 0.04±0.01 | 0.07±0.04 | 0.12±0.08 | 0.026±0.005 | 2.8±0.7 | 11.4±4.0 |
| Spleen | 0.06±0.02 | 1.02±0.27 | 1.33±0.15 | 0.19±0.05 | 0.016±0.002 | 9.4±1.2 | 4.6±0.5 |
| Thyroid | 0.07±0.06 | 1.30±0.34 | 3.86±1.19 | 0.76±0.27 | 0.024±0.002 | 11.1±3.6 | 1.6±1.5 |
| Lumbar Vertebr. Marrow | 0.01±0.01 | 0.12±0.03 | 0.30±0.08 | 0.18±0.04 | 0.023±0.002 | 3.6±0.8 | 2.8±1.1 |
| Iliac Marrow | 0.00±0.00 | 0.08±0.02 | 0.21±0.06 | 0.18±0.05 | 0.026±0.002 | 3.1±0.5 | 6.8±1.1 |
| Parotid Salivary Glands | 0.01±0.01 | 0.35±0.09 | 2.38±0.51 | 0.94±0.24 | 0.013±0.002 | 10.9±3.8 | 2.4±0.9 |
| Choroid Plexus | 0.03±0.00 | 0.06±0.03 | 4.27±1.64 | 0.46±0.12 | 0.014±0.001 | 0.5±0.1 | 4.2±0.8 |
| Axillary and Pelvic LNs | 0.03±0.04 | 0.12±0.08 | 0.78±0.91 | 0.49±0.41 | 0.033±0.109 | 5.8±7.3 | 7.4±5.3 |
| Underarm Skin | 0.01±0.01 | 0.05±0.04 | 0.65±0.83 | 0.55±0.55 | 0.022±0.007 | 3.0±1.9 | 1.2±5.9 |
| Joints | 0.00±0.01 | 0.03±0.03 | 0.71±0.43 | 0.28±0.11 | 0.029±0.009 | 1.3±1.2 | 31.7±13.7 |
| LLL Tumor Sub-Regions | 0.10±0.06 | 0.95±0.60 | 3.11±1.84 | 0.31±0.05 | 0.022±0.003 | 4.9±1.3 | 7.3±4.0 |
| Mediastinal LNs | 0.08±0.06 | 0.44±0.29 | 1.23±1.06 | 0.34±0.17 | 0.021±0.004 | 7.5±2.1 | -3.0±4.0 |

**SUPPLEMENTAL TABLE 2.** Estimated RMSE (%) of AIC-preferred model microparameters in analyzed organs-of-interest, calculated from model fitting on 90-min data from 100 simulated TACs generated for each subject from the calculated noise model of the measured TAC. The results show the averaged values among all subjects with 90-min dynamic scans. Results within <±10%, <±30%, and >±30% are marked with green, yellow, and red shadings, respectively.

| | $v_b$ | $K_1$ | $k_2$ | $k_3$ | $k_4$ | $V_T$ | $t_d$ |
|---|---|---|---|---|---|---|---|
| LV Myocardium | 7.3 | 16.2 | 39.9 | 17.5 | 11.1 | 5.4 | 15.2 |
| RV Myocardium | 3.3 | 7.9 | 6.5 | NA | NA | 15.2 | 0.0 |
| Lungs | 9.6 | 77.6 | 93.9 | 42.5 | 106.9 | 5.3 | 6.2 |
| Liver | 70.0 | 8.7 | 354.7 | 216.8 | 114.7 | 4.2 | 21.9 |
| Muscle | 49.3 | 25.0 | 258.5 | 159.1 | 70.2 | 29.4 | 59.5 |
| Spleen | 44.1 | 8.2 | 11.1 | 5.8 | 3.9 | 4.3 | 20.1 |
| Thyroid | 42.3 | 19.0 | 23.6 | 13.2 | 8.7 | 4.8 | 58.8 |
| Lumbar Vertebrae Marrow | 180.5 | 10.0 | 28.0 | 20.3 | 10.1 | 4.5 | 83.0 |
| Iliac Marrow | 200.8 | 11.7 | 46.1 | 31.9 | 12.8 | 7.7 | 48.3 |
| Parotid Salivary Glands | 211.9 | 28.6 | 46.1 | 19.7 | 37.5 | 4.7 | 109.2 |
| Choroid Plexus | 24.8 | 85.2 | 67.7 | 95.7 | 273.7 | 10.3 | 22.1 |
| Axillary and Pelvic LNs | 53.5 | 19.8 | 72.6 | 40.8 | 35.2 | 5.9 | 31.5 |
| Underarm Skin | 52.0 | 10.1 | 39.8 | 25.2 | 16.9 | 3.8 | 64.5 |
| Joints | 27.2 | 8.9 | 32.9 | 17.0 | 8.8 | 3.4 | 4.2 |
| LLL Tumor Sub-Regions | 16.0 | 14.2 | 18.7 | 11.0 | 4.7 | 3.4 | 9.3 |
| Mediastinal LNs | 30.2 | 14.5 | 28.3 | 14.3 | 8.1 | 4.5 | 29.8 |

**SUPPLEMENTAL TABLE 3.** Estimated bias (%) of AIC-preferred model microparameters in analyzed organs-of-interest, calculated from model fitting with a fixed time delay parameter on 90-min data from 100 simulated TACs generated for each subject from the calculated noise model of the measured TAC. The results show the averaged values among all subjects with 90-min dynamic scans. Results within <±10%, <±30%, and >±30% are marked with green, yellow, and red shadings, respectively.

| | $v_b$ | $K_1$ | $k_2$ | $k_3$ | $k_4$ | $V_T$ |
|---|---|---|---|---|---|---|
| LV Myocardium | 0.0 | -0.2 | -0.3 | -0.4 | 0.4 | 0.4 |
| RV Myocardium | 0.2 | 0.8 | -0.2 | NA | NA | -3.8 |
| Lungs | 0.1 | 0.8 | 2.3 | -0.4 | 1.9 | 0.5 |
| Liver | 12.5 | -0.1 | 86.5 | 7.8 | -21.4 | 1.2 |
| Muscle | -0.1 | 0.1 | 0.5 | 0.3 | -0.2 | 0.0 |
| Spleen | -0.7 | 0.2 | 0.4 | 0.1 | 0.1 | -1.3 |
| Thyroid | 0.8 | -0.3 | -0.3 | -0.1 | 0.2 | 0.1 |
| Lumbar Vertebrae Marrow | 2.5 | 0.2 | 0.7 | 0.3 | 0.0 | -0.7 |
| Iliac Marrow | 2.2 | -0.1 | -0.3 | -0.6 | 0.0 | -0.1 |
| Parotid Salivary Glands | 1.2 | -0.4 | -0.5 | -0.1 | 0.7 | 0.2 |
| Choroid Plexus | 0.5 | -3.3 | -3.5 | 0.1 | 0.7 | 1.2 |
| Axillary and Pelvic LNs | 10.2 | 3.1 | 31.1 | 17.3 | 5.6 | 0.6 |
| Underarm Skin | 3.6 | 0.2 | 3.4 | 1.0 | 1.2 | 0.5 |
| Joints | -1.1 | 1.6 | 10.5 | 3.1 | 0.1 | -0.2 |
| LLL Tumor Sub-Regions | 1.3 | -0.9 | -0.7 | -0.1 | -0.1 | 0.0 |
| Mediastinal LNs | 18.1 | 2.7 | 6.1 | 0.5 | -0.2 | 0.6 |

**SUPPLEMENTAL TABLE 4.** Estimated standard deviation (%) of AIC-preferred model microparameters in analyzed organs-of-interest, calculated from model fitting with a fixed time delay parameter on 90-min data from 100 simulated TACs generated for each subject from the calculated noise model of the measured TAC. The results show the averaged values among all subjects with 90-min dynamic scans. Results within <±10%, <±30%, and >±30% are marked with green, yellow, and red shadings, respectively.

| | $v_b$ | $K_1$ | $k_2$ | $k_3$ | $k_4$ | $V_T$ |
|---|---|---|---|---|---|---|
| LV Myocardium | 2.6 | 5.4 | 12.5 | 6.4 | 3.9 | 1.5 |
| RV Myocardium | 2.7 | 6.3 | 5.0 | NA | NA | 8.3 |
| Lungs | 3.2 | 25.8 | 38.4 | 14.5 | 10.0 | 1.1 |
| Liver | 113.0 | 1.2 | 35.8 | 67.6 | 22.4 | 2.9 |
| Muscle | 4.8 | 1.4 | 8.8 | 10.2 | 5.2 | 1.6 |
| Spleen | 18.4 | 5.6 | 7.9 | 4.3 | 2.9 | 1.8 |
| Thyroid | 12.7 | 8.4 | 11.6 | 5.6 | 2.7 | 1.2 |
| Lumbar Vertebrae Marrow | 31.2 | 3.8 | 10.3 | 7.7 | 3.8 | 1.5 |
| Iliac Marrow | 36.0 | 3.5 | 12.1 | 9.5 | 4.2 | 1.3 |
| Parotid Salivary Glands | 28.4 | 8.2 | 14.1 | 5.0 | 3.9 | 1.1 |
| Choroid Plexus | 4.9 | 20.5 | 22.5 | 14.3 | 6.2 | 2.3 |
| Axillary and Pelvic LNs | 49.3 | 19.4 | 82.9 | 36.0 | 28.8 | 4.3 |
| Underarm Skin | 27.2 | 8.0 | 31.5 | 20.1 | 11.5 | 2.6 |
| Joints | 20.2 | 8.8 | 40.3 | 19.7 | 10.7 | 2.5 |
| LLL Tumor Sub-Regions | 12.5 | 12.1 | 16.6 | 9.0 | 4.7 | 2.2 |
| Mediastinal LNs | 56.6 | 14.1 | 31.5 | 14.7 | 7.4 | 2.8 |

**SUPPLEMENTAL TABLE 5.** Estimated RMSE (%) of AIC-preferred model microparameters in analyzed organs-of-interest, calculated from model fitting with a fixed time delay parameter on 90-min data from 100 simulated TACs generated for each subject from the calculated noise model of the measured TAC. The results show the averaged values among all subjects with 90-min dynamic scans. Results within <±10%, <±30%, and >±30% are marked with green, yellow, and red shadings, respectively.

| | $v_b$ | $K_1$ | $k_2$ | $k_3$ | $k_4$ | $V_T$ |
|---|---|---|---|---|---|---|
| LV Myocardium | 2.6 | 5.4 | 12.6 | 6.4 | 3.9 | 2.2 |
| RV Myocardium | 2.7 | 6.3 | 5.0 | NA | NA | 10.0 |
| Lungs | 3.2 | 26.8 | 40.7 | 14.6 | 10.3 | 1.5 |
| Liver | 130.2 | 2.7 | 115.4 | 77.7 | 56.7 | 3.9 |
| Muscle | 4.8 | 1.4 | 8.9 | 10.2 | 5.2 | 2.0 |
| Spleen | 18.3 | 5.6 | 7.8 | 4.3 | 2.9 | 2.4 |
| Thyroid | 12.7 | 8.6 | 11.9 | 5.7 | 2.7 | 2.1 |
| Lumbar Vertebrae Marrow | 31.3 | 3.8 | 10.3 | 7.7 | 3.8 | 2.1 |
| Iliac Marrow | 36.1 | 3.5 | 12.1 | 9.6 | 4.2 | 2.0 |
| Parotid Salivary Glands | 28.3 | 8.2 | 14.1 | 5.0 | 4.0 | 1.3 |
| Choroid Plexus | 5.0 | 21.8 | 25.1 | 14.3 | 6.2 | 3.1 |
| Axillary and Pelvic LNs | 51.7 | 19.9 | 85.5 | 37.1 | 30.7 | 5.4 |
| Underarm Skin | 27.6 | 8.1 | 32.4 | 20.7 | 11.8 | 3.5 |
| Joints | 20.2 | 9.0 | 41.8 | 20.1 | 10.7 | 3.4 |
| LLL Tumor Sub-Regions | 12.6 | 12.6 | 17.3 | 9.0 | 4.7 | 2.8 |
| Mediastinal LNs | 61.1 | 14.4 | 32.2 | 14.7 | 7.4 | 3.8 |

**SUPPLEMENTAL TABLE 6.** Estimated bias (%) of AIC-preferred model microparameters in analyzed organs-of-interest, calculated from model fitting on 60-min data from 100 simulated TACs generated for each subject from the calculated noise model of the measured TAC. The results show the averaged values among all subjects with 90-min dynamic scans. Results within <±10%, <±30%, and >±30% are marked with green, yellow, and red shadings, respectively.

| | $v_b$ | $K_1$ | $k_2$ | $k_3$ | $k_4$ | $V_T$ | $t_d$ |
|---|---|---|---|---|---|---|---|
| LV Myocardium | 0.3 | -0.4 | 1.6 | -0.9 | 1.5 | 1.6 | 1.4 |
| RV Myocardium | 0.0 | 0.7 | -0.1 | NA | NA | 6.7 | 0.0 |
| Lungs | 0.2 | 12.3 | 12.1 | -2.9 | 65.7 | 2.1 | 0.0 |
| Liver | 30.7 | 1.9 | 175.0 | 110.2 | 82.1 | 2.1 | 15.3 |
| Muscle | 7.6 | 9.2 | 223.2 | 242.9 | 32.9 | 0.0 | 26.9 |
| Spleen | -0.2 | -0.5 | -0.4 | -0.2 | -0.1 | -1.3 | -1.3 |
| Thyroid | 9.8 | -5.0 | -5.5 | 1.7 | 2.1 | 1.7 | 25.8 |
| Lumbar Vertebrae Marrow | 95.8 | 0.3 | 1.8 | 0.3 | -0.5 | 5.8 | 18.4 |
| Iliac Marrow | 112.2 | 1.8 | 10.7 | 5.9 | -0.5 | 4.4 | 18.2 |
| Parotid Salivary Glands | 104.5 | -12.1 | -20.9 | -5.6 | 15.8 | 3.1 | 39.9 |
| Choroid Plexus | 2.2 | -7.5 | -27.6 | 9.6 | 240.5 | 0.0 | -0.9 |
| Axillary and Pelvic LNs | 16.5 | 1.0 | 35.7 | 6.4 | 10.9 | 2.3 | -0.8 |
| Underarm Skin | 25.1 | -2.5 | 1.0 | 2.0 | 4.6 | 3.7 | -16.8 |
| Joints | -0.2 | 2.4 | 11.2 | 3.0 | -0.4 | 1.7 | -0.4 |
| LLL Tumor Sub-Regions | 0.4 | -1.2 | -0.9 | 0.5 | 0.3 | -0.8 | -0.7 |
| Mediastinal LNs | 94.7 | 1.0 | 3.4 | 0.8 | 0.7 | 0.3 | -2.7 |

**SUPPLEMENTAL TABLE 7.** Estimated standard deviation (%) of AIC-preferred model microparameters in analyzed organs-of-interest, calculated from model fitting on 60-min data from 100 simulated TACs generated for each subject from the calculated noise model of the measured TAC. The results show the averaged values among all subjects with 90-min dynamic scans. Results within <±5%, <±30%, and >±30% are marked with green, yellow, and red shadings, respectively.

| | $v_b$ | $K_1$ | $k_2$ | $k_3$ | $k_4$ | $V_T$ | $t_d$ |
|---|---|---|---|---|---|---|---|
| LV Myocardium | 7.3 | 17.7 | 41.2 | 17.8 | 12.5 | 5.7 | -13.6 |
| RV Myocardium | 3.1 | 7.4 | 10.7 | NA | NA | 15.4 | 0.0 |
| Lungs | 9.4 | 64.7 | 83.0 | 49.8 | 303.8 | 6.0 | 6.7 |
| Liver | 64.3 | 3.3 | 150.4 | 147.6 | 129.6 | 4.9 | 15.3 |
| Muscle | 39.7 | 25.7 | 330.6 | 231.9 | 215.6 | 0.0 | 48.6 |
| Spleen | 43.3 | 7.8 | 10.7 | 5.8 | 5.5 | 4.3 | 19.8 |
| Thyroid | 38.0 | 16.5 | 20.8 | 12.2 | 7.2 | 3.5 | 48.5 |
| Lumbar Vertebrae Marrow | 144.3 | 10.0 | 27.9 | 21.2 | 14.5 | 7.7 | 75.2 |
| Iliac Marrow | 172.2 | 11.9 | 47.9 | 37.2 | 19.2 | 12.0 | 51.8 |
| Parotid Salivary Glands | 160.2 | 22.8 | 35.9 | 18.0 | 28.5 | 7.3 | 87.6 |
| Choroid Plexus | 23.9 | 78.5 | 59.1 | 97.1 | 483.5 | 0.0 | 22.4 |
| Axillary and Pelvic LNs | 49.1 | 21.1 | 74.9 | 41.5 | 43.6 | 9.4 | 17.0 |
| Underarm Skin | 49.4 | 7.9 | 36.6 | 26.2 | 16.9 | 7.8 | -3.9 |
| Joints | 26.3 | 8.9 | 38.0 | 20.4 | 11.6 | 5.5 | 4.2 |
| LLL Tumor Sub-Regions | 16.9 | 13.8 | 17.8 | 11.1 | 6.3 | 3.7 | 9.9 |
| Mediastinal LNs | 122.1 | 15.3 | 31.0 | 15.9 | 10.3 | 5.1 | -15.8 |

**SUPPLEMENTAL TABLE 8.** Estimated RMSE (%) of AIC-preferred model microparameters in analyzed organs-of-interest, calculated from model fitting on 60-min data from 100 simulated TACs generated for each subject from the calculated noise model of the measured TAC. The results show the averaged values among all subjects with 90-min dynamic scans. Results within <±5%, <±30%, and >±30% are marked with green, yellow, and red shadings, respectively.

| | $v_b$ | $K_1$ | $k_2$ | $k_3$ | $k_4$ | $V_T$ | $t_d$ |
|---|---|---|---|---|---|---|---|
| LV Myocardium | 7.3 | 18.1 | 42.1 | 17.7 | 12.7 | 6.4 | 13.6 |
| RV Myocardium | 3.1 | 7.4 | 10.7 | NA | NA | 23.0 | 0.0 |
| Lungs | 9.4 | 67.4 | 89.5 | 50.1 | 309.4 | 9.0 | 6.7 |
| Liver | 71.0 | 7.3 | 258.0 | 224.2 | 204.0 | 5.4 | 21.9 |
| Muscle | 40.2 | 27.2 | 345.7 | 247.9 | 218.4 | 0.0 | 57.6 |
| Spleen | 43.2 | 7.8 | 10.7 | 5.8 | 5.5 | 6.6 | 19.8 |
| Thyroid | 39.6 | 17.6 | 22.2 | 12.7 | 7.5 | 4.4 | 58.7 |
| Lumbar Vertebrae Marrow | 179.9 | 10.0 | 27.9 | 21.2 | 14.5 | 9.7 | 83.2 |
| Iliac Marrow | 210.4 | 12.0 | 49.0 | 37.6 | 19.2 | 15.6 | 55.5 |
| Parotid Salivary Glands | 193.4 | 26.3 | 42.9 | 19.7 | 32.8 | 12.3 | 100.0 |
| Choroid Plexus | 24.0 | 82.6 | 72.6 | 97.2 | 488.6 | 0.0 | 22.3 |
| Axillary and Pelvic LNs | 54.4 | 22.0 | 78.7 | 44.0 | 47.2 | 11.8 | 32.2 |
| Underarm Skin | 56.9 | 9.6 | 41.8 | 28.2 | 19.5 | 9.5 | 65.6 |
| Joints | 26.3 | 9.3 | 39.8 | 20.7 | 11.6 | 2.5 | 6.8 |
| LLL Tumor Sub-Regions | 17.0 | 14.5 | 18.6 | 11.2 | 6.3 | 4.9 | 10.3 |
| Mediastinal LNs | 30.1 | 15.6 | 31.4 | 16.1 | 10.4 | 6.2 | 31.9 |

### Supplementary Figures

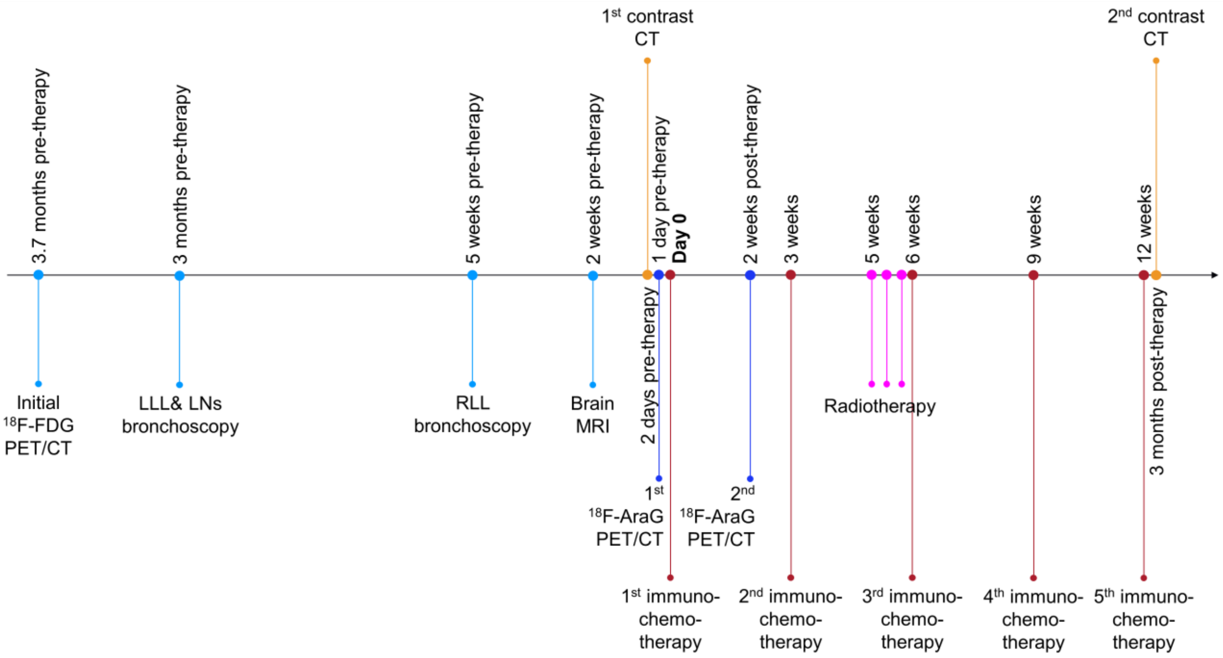


**SUPPLEMENTAL FIGURE 1.** Graphical timeline of the relevant medical interventions in the NSCLC patient (Sub05). Patient had an $^{18}$F-FDG PET/CT scan, which showed bilateral pulmonary masses with several FDG-avid mediastinal and left hilar lymph nodes. Patient underwent bronchoscopy of the left lower lobe (LLL) mass and mediastinal LNs. Bronchoscopy of LLL mass showed adenocarcinoma, PD-L1<1%, One of three lymph nodes (station 4L) was found to be suspicious for metastatic carcinoma. After discussion with the Thoracic Multidisciplinary Tumor Board, the recommended plan was to pursue right lower lobe (RLL) biopsy, which revealed non-small cell carcinoma with glandular features, double primary. Patient had a brain MRI scan, which showed no evidence of metastatic disease. Patient was enrolled in a clinical trial for a randomized phase II/III trial of modern immunotherapy based systemic therapy with or without stereotactic body radiation therapy for PD-L1-negative, advanced NSCLC. A contrast-enhanced chest CT scan and an initial $^{18}$F-AraG PET/CT scan were performed 2 days and 1 day before the therapy start, respectively. The patient was started on anti-PD-1 immunotherapy (pembrolizumab) combined with chemotherapy (pemetrexed-carboplatin) every three weeks. Before the third immunotherapy cycle, the patient received a course of stereotactic body radiation therapy to the LLL mass with palliative intent, with a total dose of 2,400 cGy given in three fractions. The second contrast-enhanced chest CT scan was performed after 5 cycles of immunotherapy.

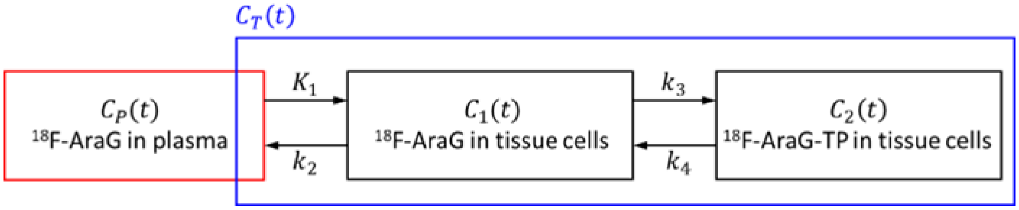


**SUPPLEMENTAL FIGURE 2.** A reversible two-tissue (2T) compartmental model, describing the kinetics of $^{18}$F-AraG in tissue. The 2T model consists of five microparameters of ($\boldsymbol{K_1}, \boldsymbol{k_2}, \boldsymbol{k_3}, \boldsymbol{k_4}, \boldsymbol{v_b}$), which will be estimated during the model fitting. $\boldsymbol{C_T(t)}$ is the activity concentration of the tissue-of-interest measured on dynamic PET images as a function of time, $\boldsymbol{t}$. $\boldsymbol{C_P(t)}$ is the activity concentration of $^{18}$F-AraG in arterial blood plasma and the input function (IF) to the system, either measured by arterial blood sampling during the dynamic scan or estimated from measurements of whole-blood activity concentration on dynamic PET images in certain blood-pool regions (e.g., left ventricle (LV), right ventricle (RV), descending aorta, ascending aorta, etc.). $\boldsymbol{C_1(t)}$ and $\boldsymbol{C_2(t)}$ are the two tissue compartments, representing concentrations of free $^{18}$F-AraG and tri-phosphorylated $^{18}$F-AraG ($^{18}$F-AraG-TP) in tissue cells, respectively. $\boldsymbol{K_1}$ (mL/min/$mL_{Tissue}$) and $\boldsymbol{k_2}$ ($min^{-1}$) are the rates of $^{18}$F-AraG transport between arterial blood plasma and tissue cells and are blood-flow de. $\boldsymbol{k_3}$ ($min^{-1}$) and $\boldsymbol{k_4}$ ($min^{-1}$) are $^{18}$F-AraG phosphorylation and dephosphorylation rates, respectively and $\boldsymbol{v_b}$ is the fractional blood volume in tissue. The irreversible 2T compartmental model ($\boldsymbol{k_4 = 0}$) and the 1T compartmental model ($\boldsymbol{k_3 = 0}$ and $\boldsymbol{k_4 = 0}$) can be considered as special cases of the reversible 2T compartmental model.

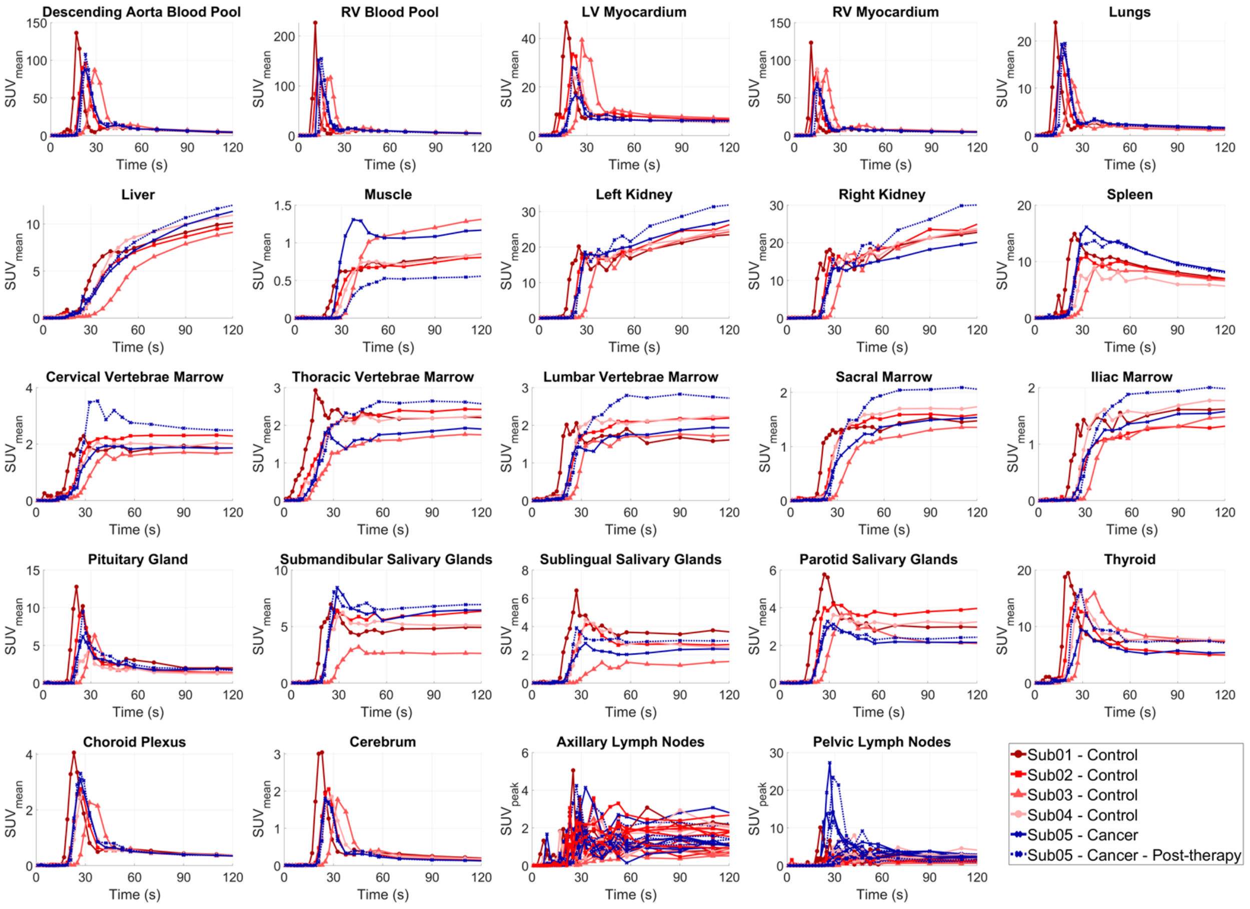


**SUPPLEMENTAL FIGURE 3.** TACs of the healthy organs-of-interest analyzed in all subjects, zoomed in on the first 2 min of the dynamic scans.

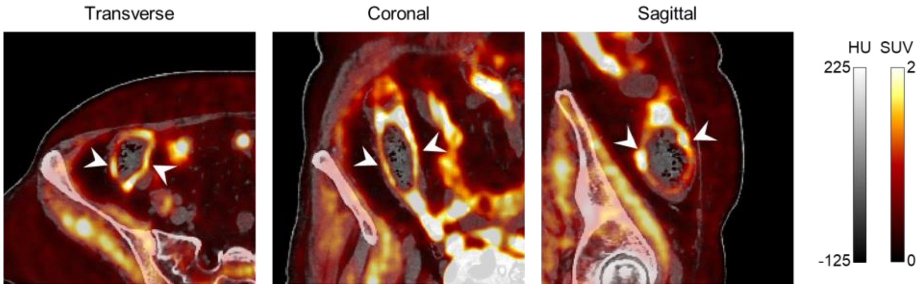


**SUPPLEMENTAL FIGURE 4.** Transverse, coronal, and sagittal PET/CT image slices of the Sub03 (50–60 min p.i.) showing uptake in the subject's ascending colon wall.

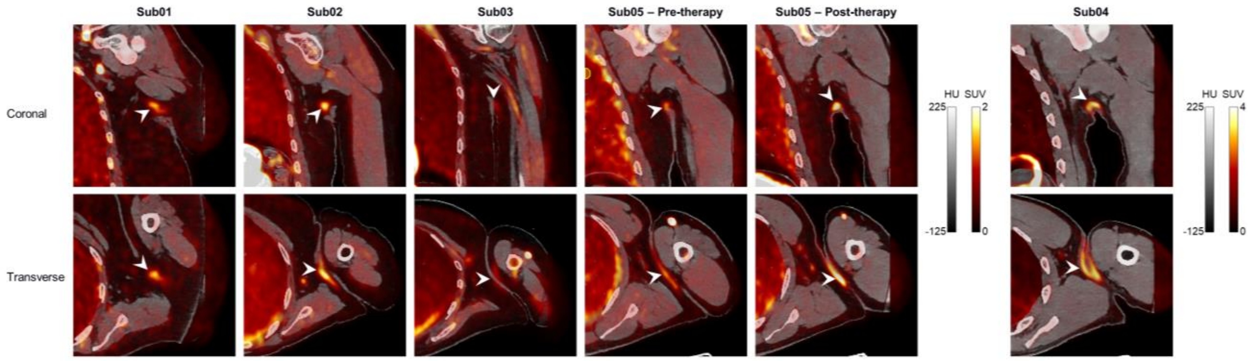


**SUPPLEMENTAL FIGURE 5.** Example coronal and transverse PET/CT slices (50–60 min p.i.) of the left underarm region in all subjects showing increased uptake in the underarm skin of all subjects compared to other skin regions. Images of Sub04 are shown on a different color scale due to the significantly higher uptake compared to other subjects.

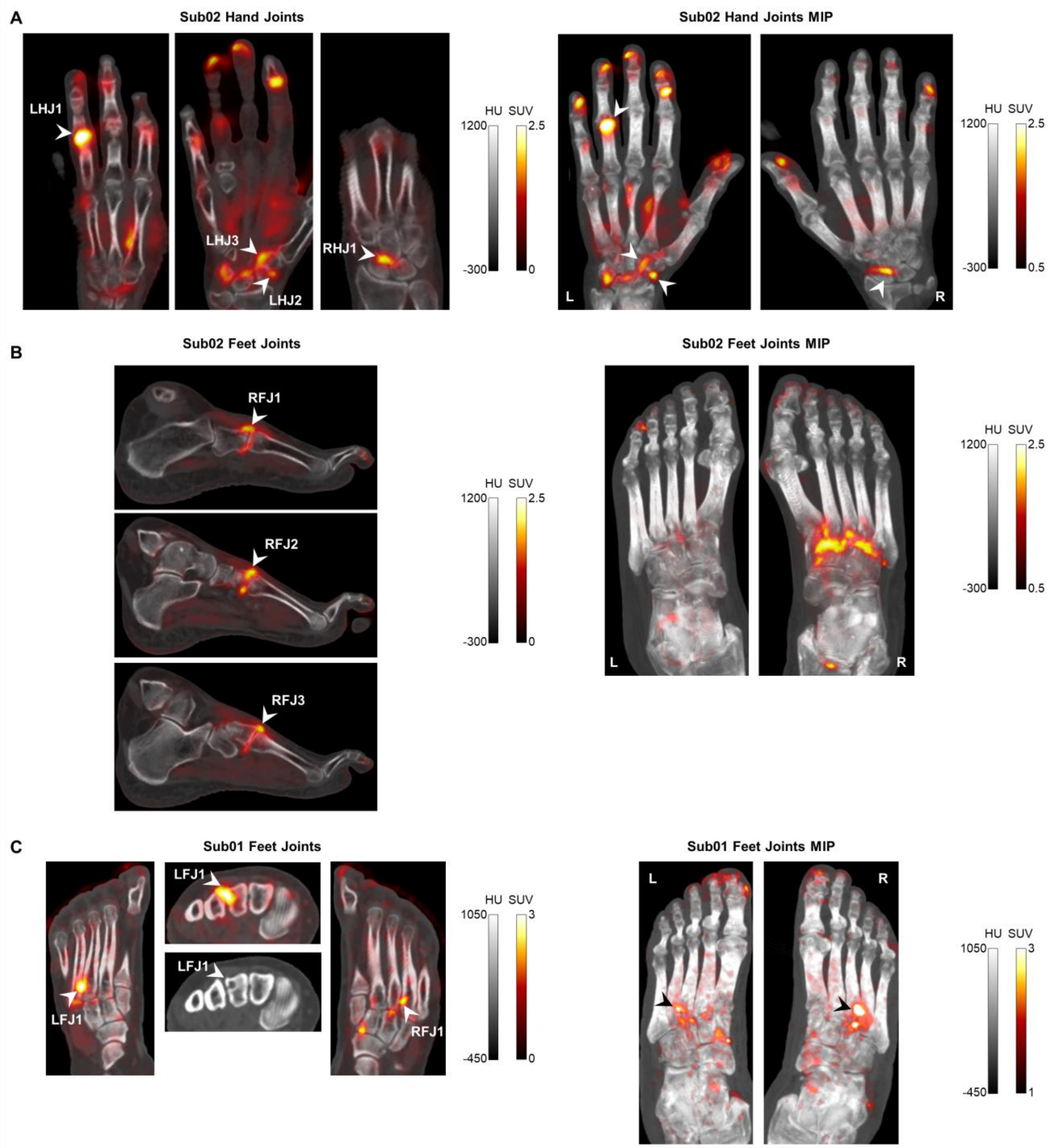


**SUPPLEMENTAL FIGURE 6.** $^{18}$F-AraG PET/CT slices (left) and maximum intensity projections (right) of the 50–60 min p.i. images of the analyzed regions of (A) hands and (B) feet joints in Sub02 who is previously diagnose with osteoarthritis and (C) feet joints in Sub01 showing high focal uptake compared to other joint regions. The location of the analyzed VOIs have been marked with arrowheads accompanied by VOI label names used throughout the paper. Foot joints analyzed in Sub02 were 4$^{th}$, 2$^{nd}$, and 3$^{rd}$ tarsometatarsal (TMT) joints of the right foot, corresponding to RFJ1, RFJ2, and RF3 VOIs, respectively. Feet joints analyzed in Sub01 were in the 4$^{th}$ and 3$^{rd}$ TMT joints of left and right feet, respectively. The focal uptake marked in the right foot (RFJ1) of Sub01 corresponded to a subcortical cystic area visualized on the CT image. LHJ, left hand joint; RHJ, right hand joint; LFJ, left foot joint; RFJ, right foot joint.

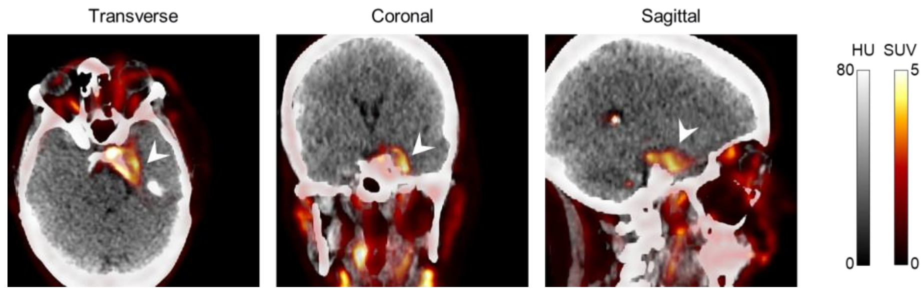


**SUPPLEMENTAL FIGURE 7.** Transverse, coronal, and sagittal PET/CT image slices (50–60 min p.i.) of Sub03 showing high uptake in the meningioma region.

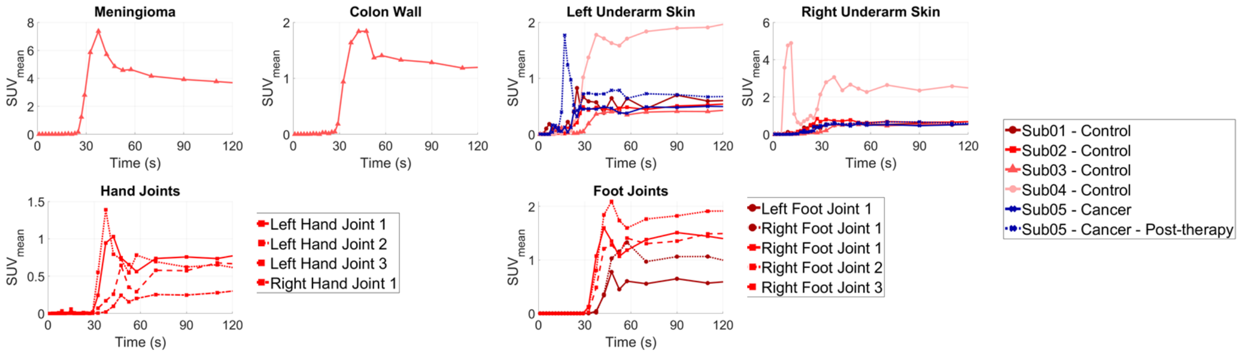


**SUPPLEMENTAL FIGURE 8.** TACs of the tissues with focally high uptake pattern in all subjects, zoomed in on the first 2 min of the dynamic scans.

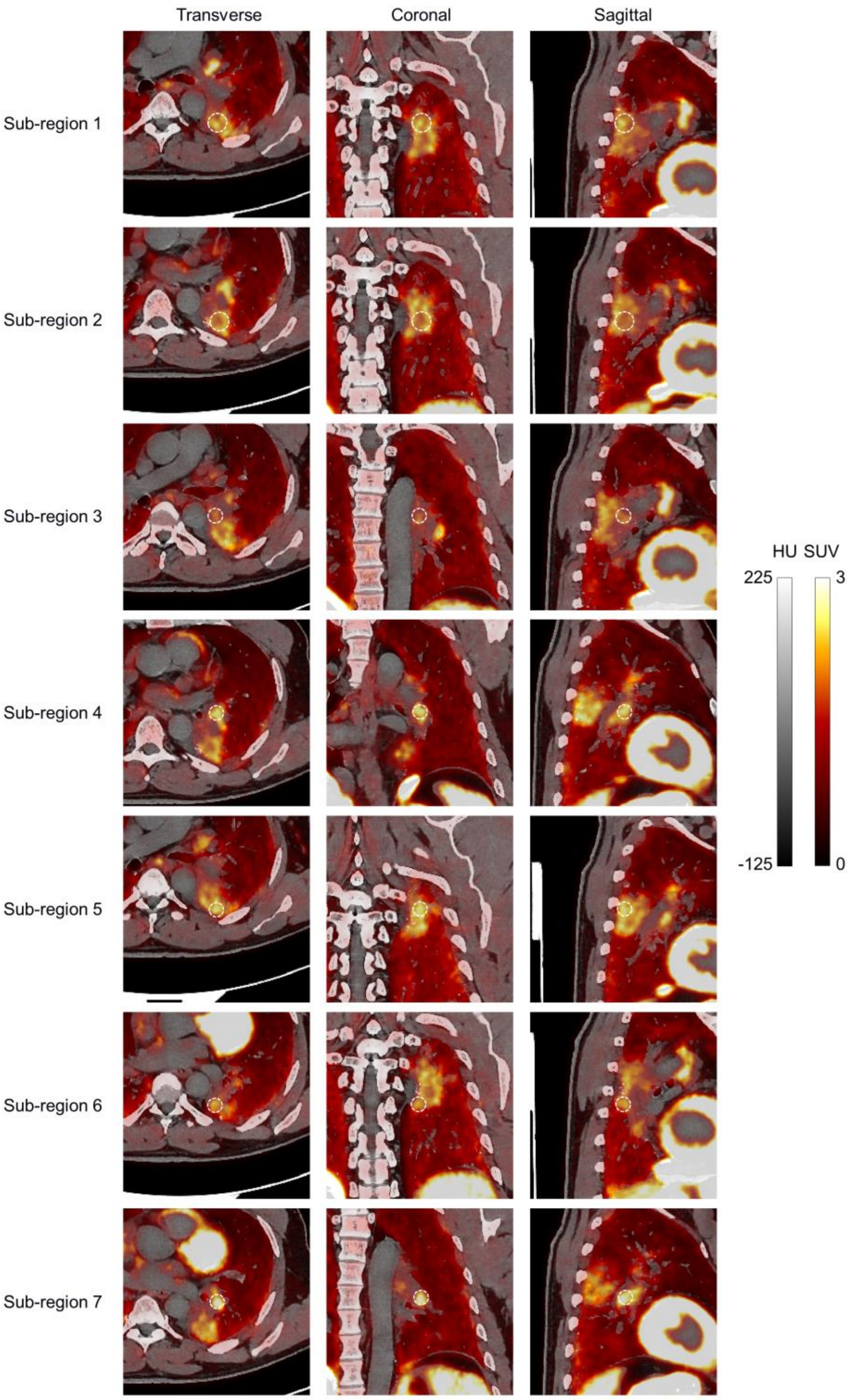


**SUPPLEMENTAL FIGURE 9.** Transverse, coronal, and sagittal PET/CT image slices (60–90 min p.i.) of the pre-therapy scan of Sub05 showing the seven analyzed sub-regions of the LLL mass. The dashed white circles show the position and dimension of the seven VOIs used for analyzing the LLL mass. VOIs 1–2 were 20 mm in diameter and VOIs 3–7 were 15 mm in diameter.

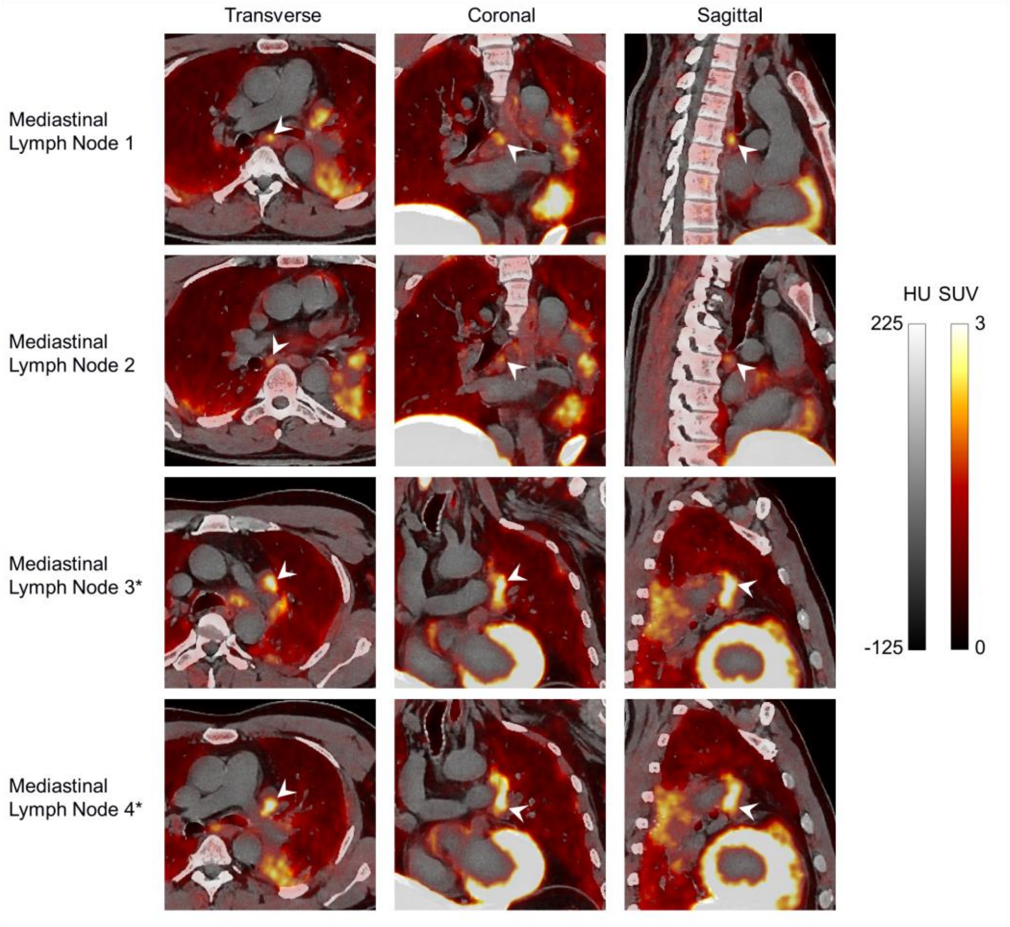


**SUPPLEMENTAL FIGURE 10.** Transverse, coronal, and sagittal PET/CT image slices (60–90 min p.i.) of the pre-therapy scan of Sub05 showing the four analyzed mediastinal LNs in proximity of the LLL mass. The two LNs marked with asterisks in their labels (3 and 4) were LNs identified as enlarged and potentially metastatic. 12-mm and 10-mm diameter spherical VOIs were used for SUVpeak calculations in non-enlarged and enlarged LNs, respectively.

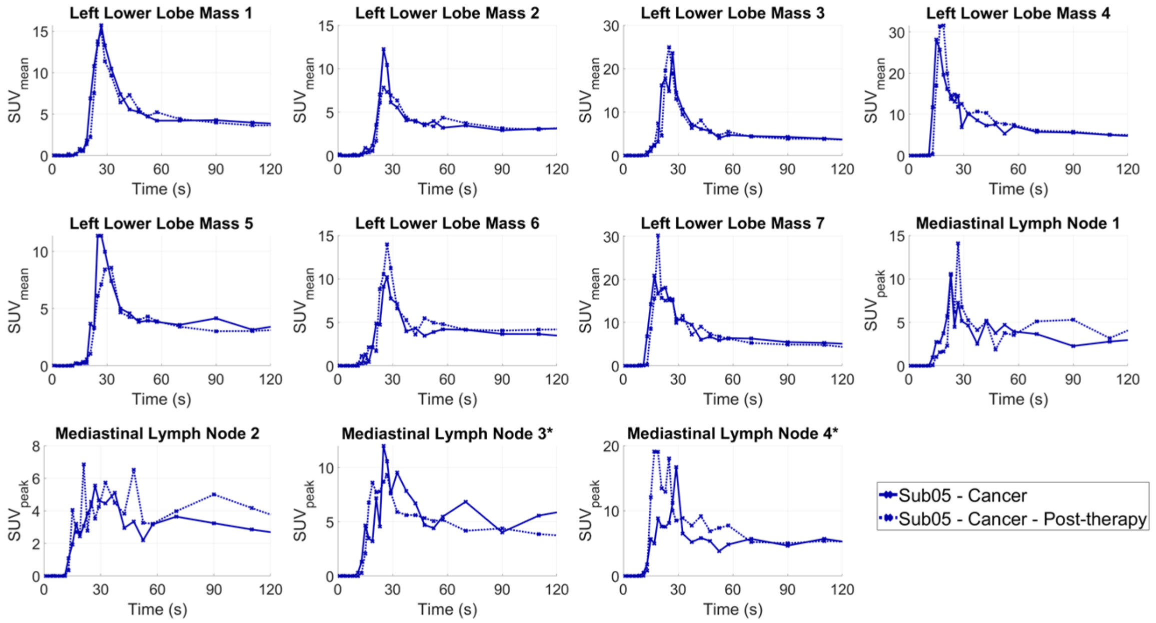

**SUPPLEMENTAL FIGURE 11.** TACs of the seven sub-regions of the LLL tumor mass and four mediastinal LNs in Sub05 pre- and post-therapy, zoomed in on the first 2 min of the dynamic scans.

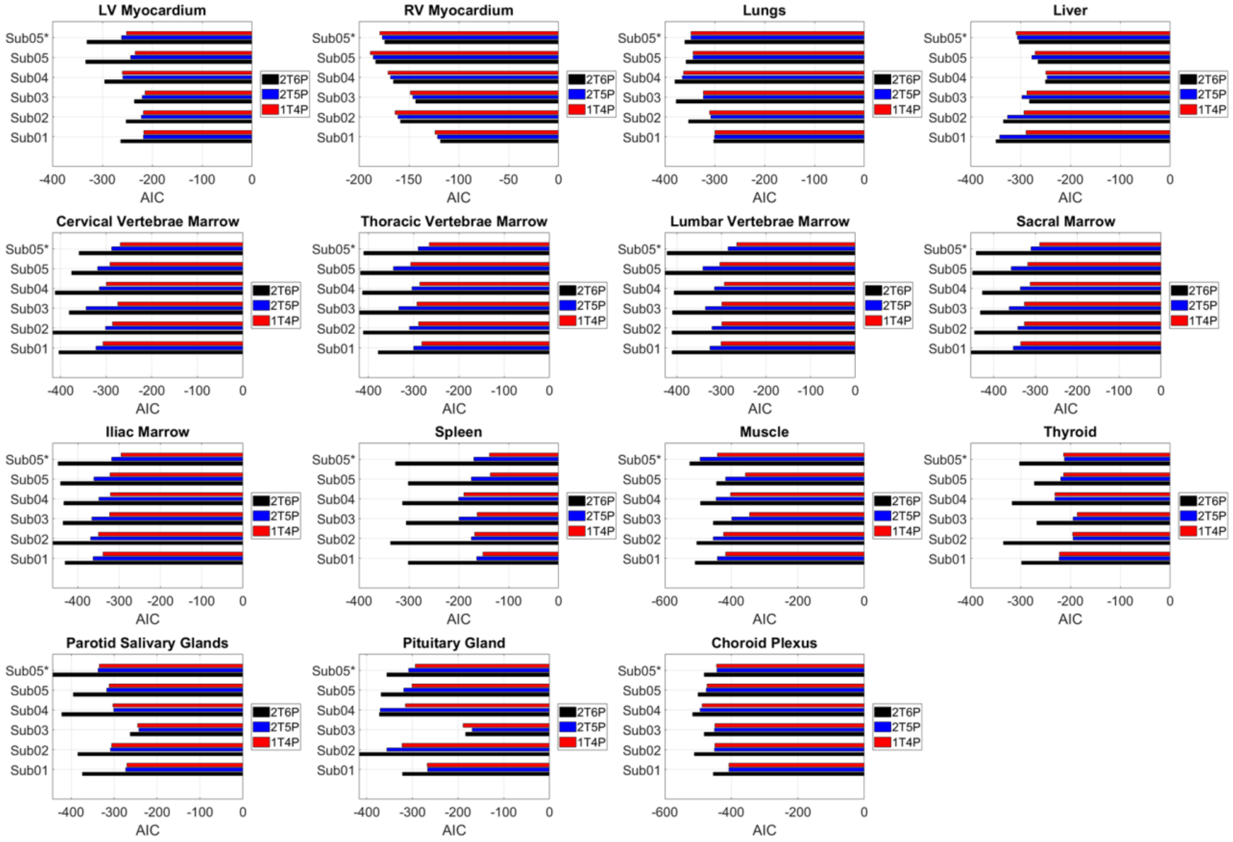

**SUPPLEMENTAL FIGURE 12.** Comparison of the AIC of the 2T6P (reversible), 2T5P (irreversible), and 1T4P model fitting in analyzed healthy organs-of-interest of all subjects. Sub05 marked with asterisk represents the post-therapy scan of Sub05.

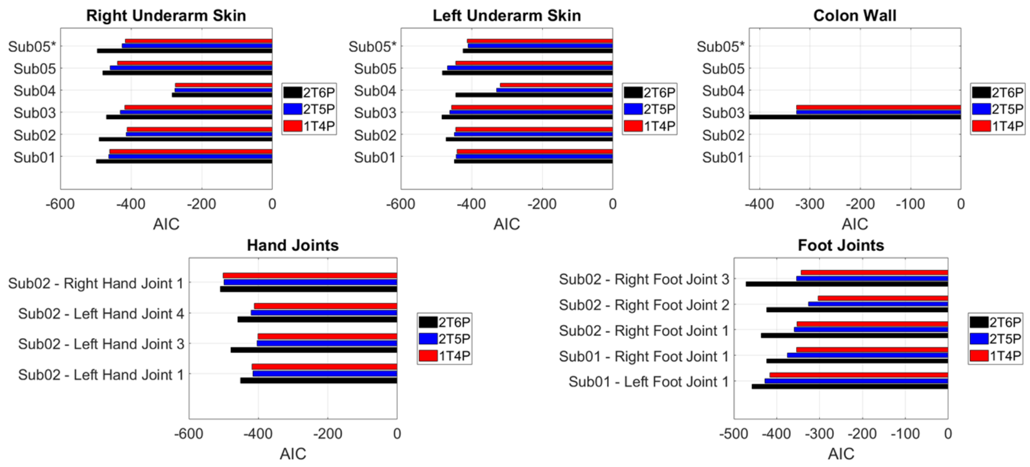


**SUPPLEMENTAL FIGURE 13.** Comparison of the AIC of the 2T6P (reversible), 2T5P (irreversible), and 1T4P model fitting in the organs with focally high uptake pattern in all subjects.

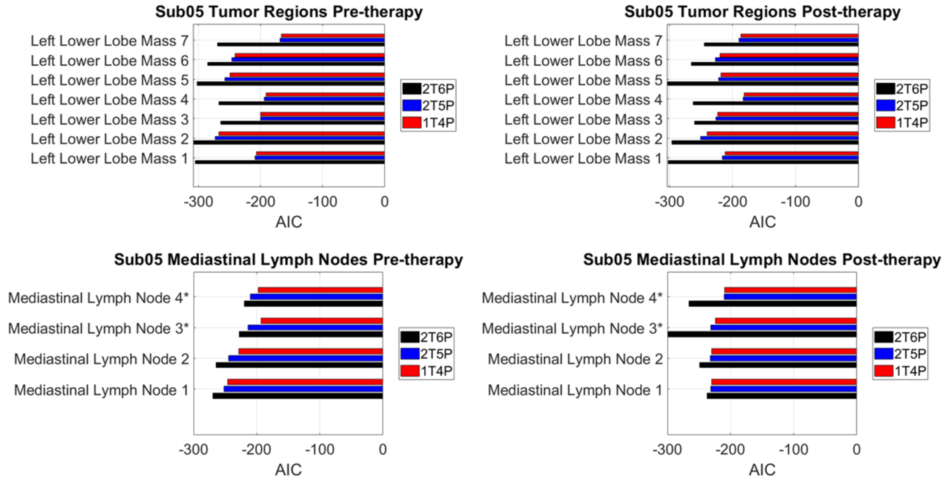


**SUPPLEMENTAL FIGURE 14.** Comparison of the AIC of the 2T6P (reversible), 2T5P (irreversible), and 1T4P model fitting in seven sub-regions of the LLL tumor mass and four mediastinal LNs in Sub05 pre- and post-therapy. LNs marked with asterisks represent enlarged LNs.

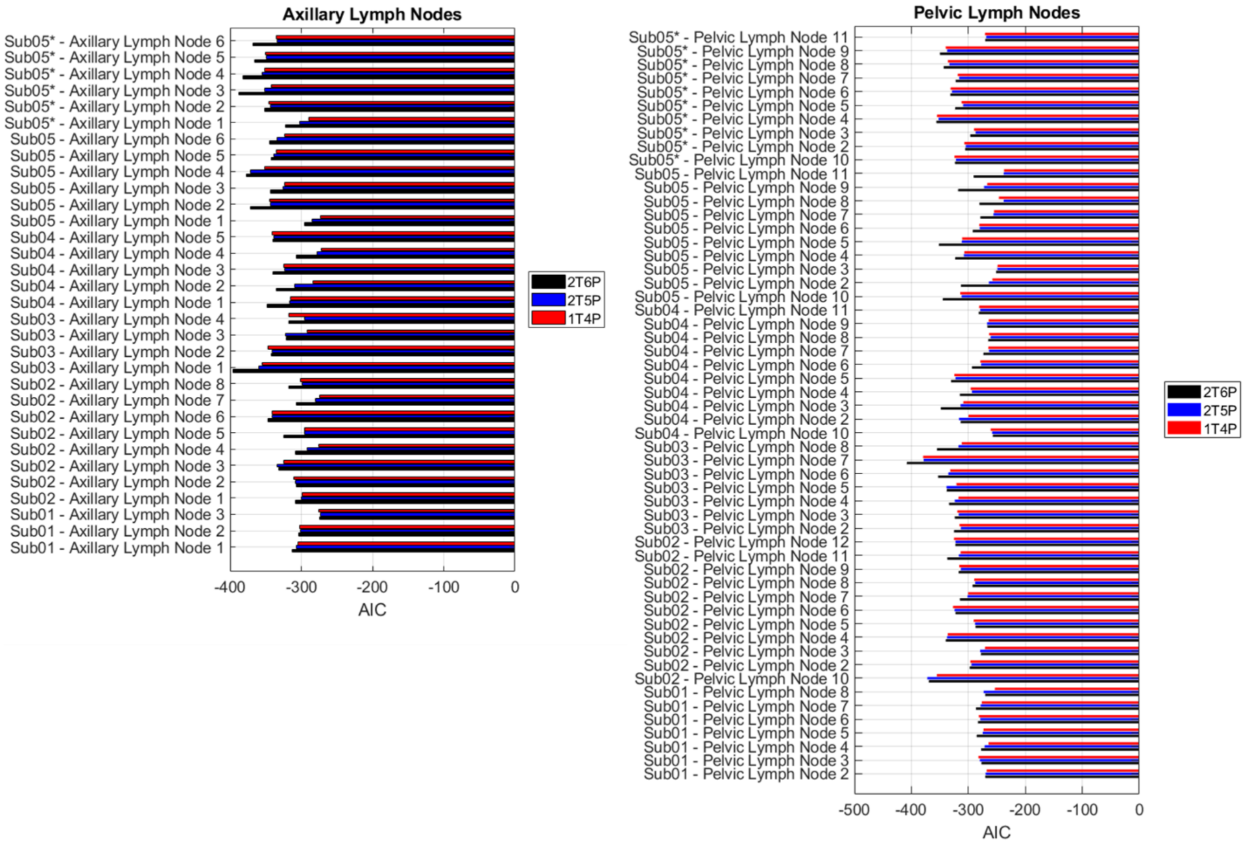


**SUPPLEMENTAL FIGURE 15.** Comparison of the AIC of the 2T6P (reversible), 2T5P (irreversible), and 1T4P model fitting in 93 total analyzed axillary and pelvic LNs in all subjects. Sub05 marked with asterisk represents the post-therapy scan of Sub05.

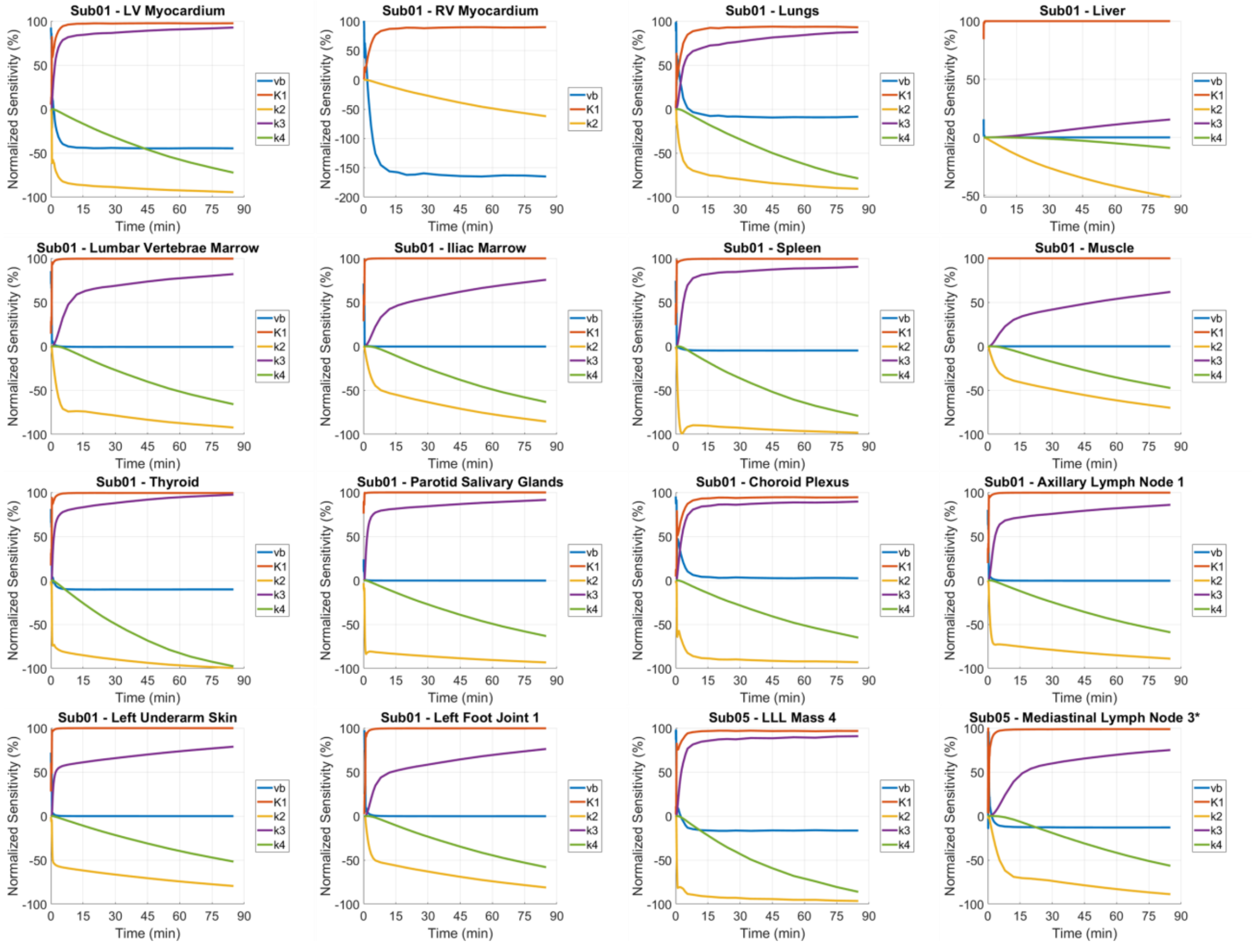


**SUPPLEMENTAL FIGURE 16.** Example normalized sensitivity plots of different organs-of-interest, comparing the normalized sensitivities of the AIC-preferred model microparameters.

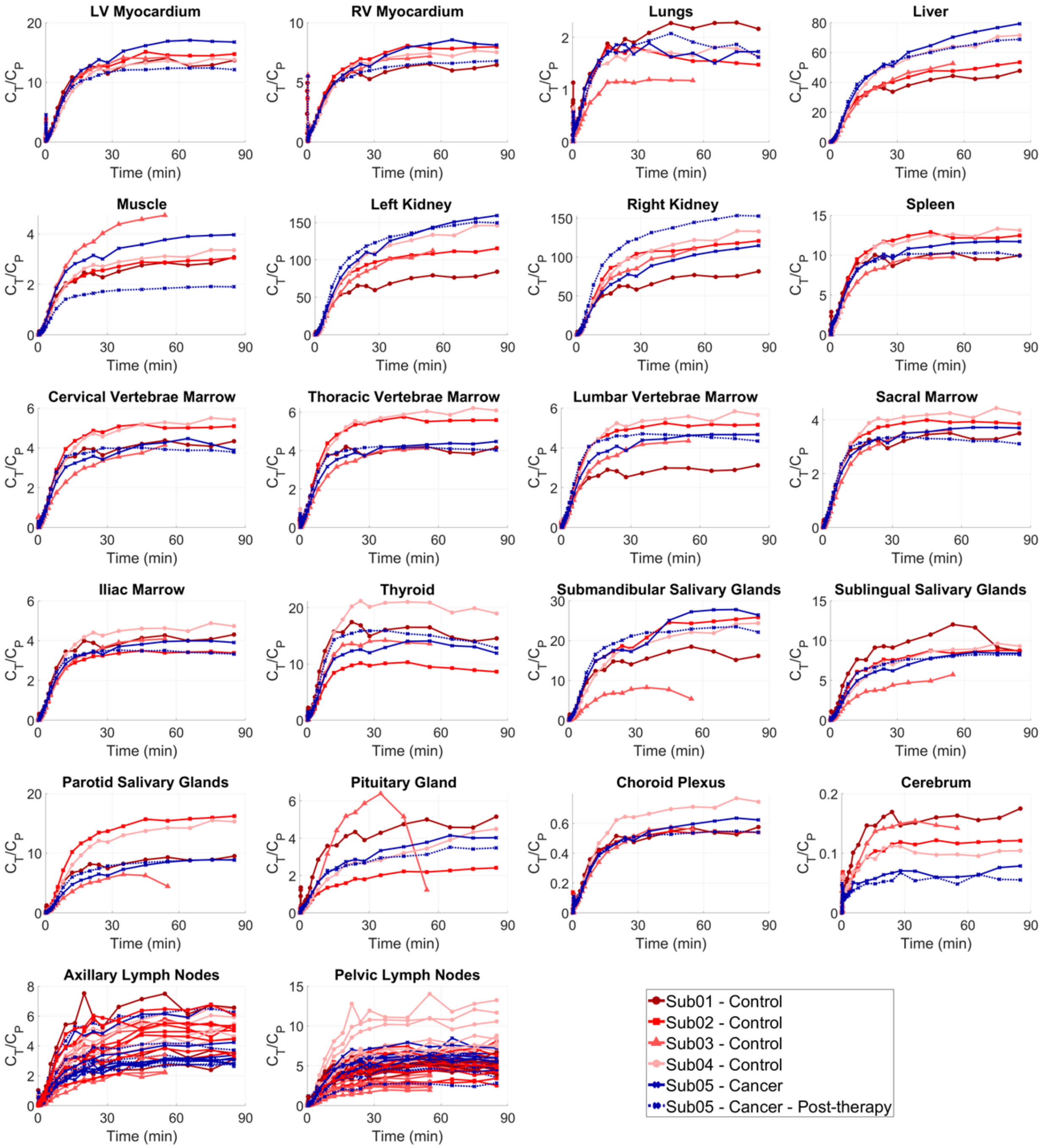


**SUPPLEMENTAL FIGURE 17.** SUVR curves of healthy organs-of-interest analyzed in all subjects plotted as a function of time during the 90 min dynamic scans, using the original blood-pool TACs. The last two datapoints in the SUVR curves in the head region of Sub03 were affected by subject's head motion.

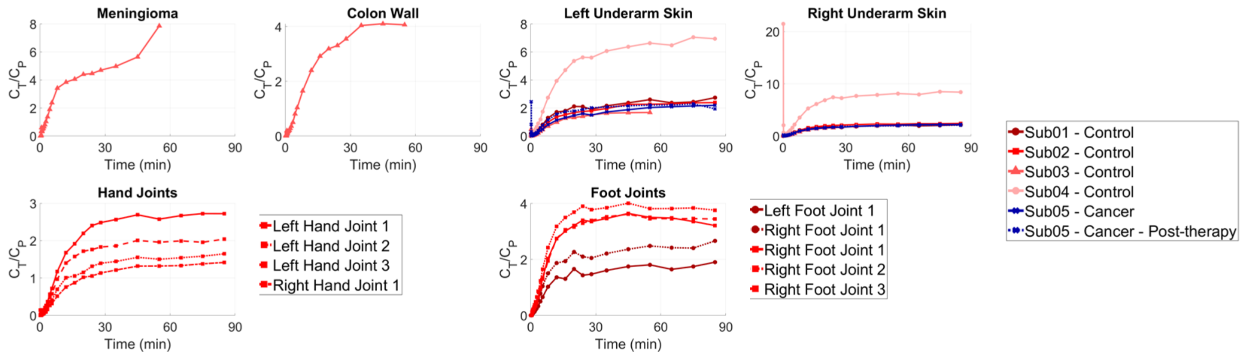


**SUPPLEMENTAL FIGURE 18.** SUVR curves of organs with focally high uptake pattern in all subjects plotted as a function of time during the 90 min dynamic scans, using the original blood-pool TACs. The last two datapoints in the SUVR curves in the head region of Sub03 were affected by subject's head motion.

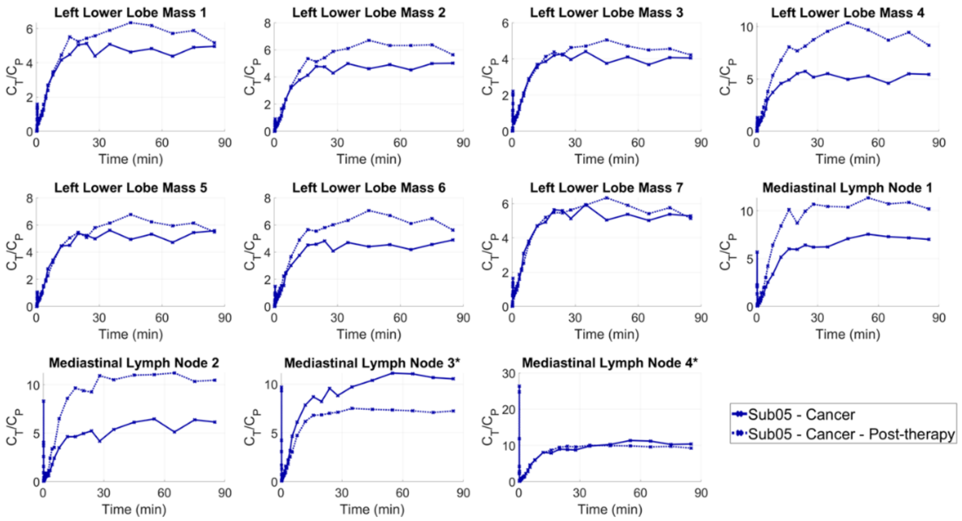


**SUPPLEMENTAL FIGURE 19.** SUVR curves of seven sub-regions of the LLL mass and the four mediastinal LNs, plotted as a function of time during the 90 min dynamic scans, using the original blood-pool TACs. The two enlarged LNs are marked with asterisks.

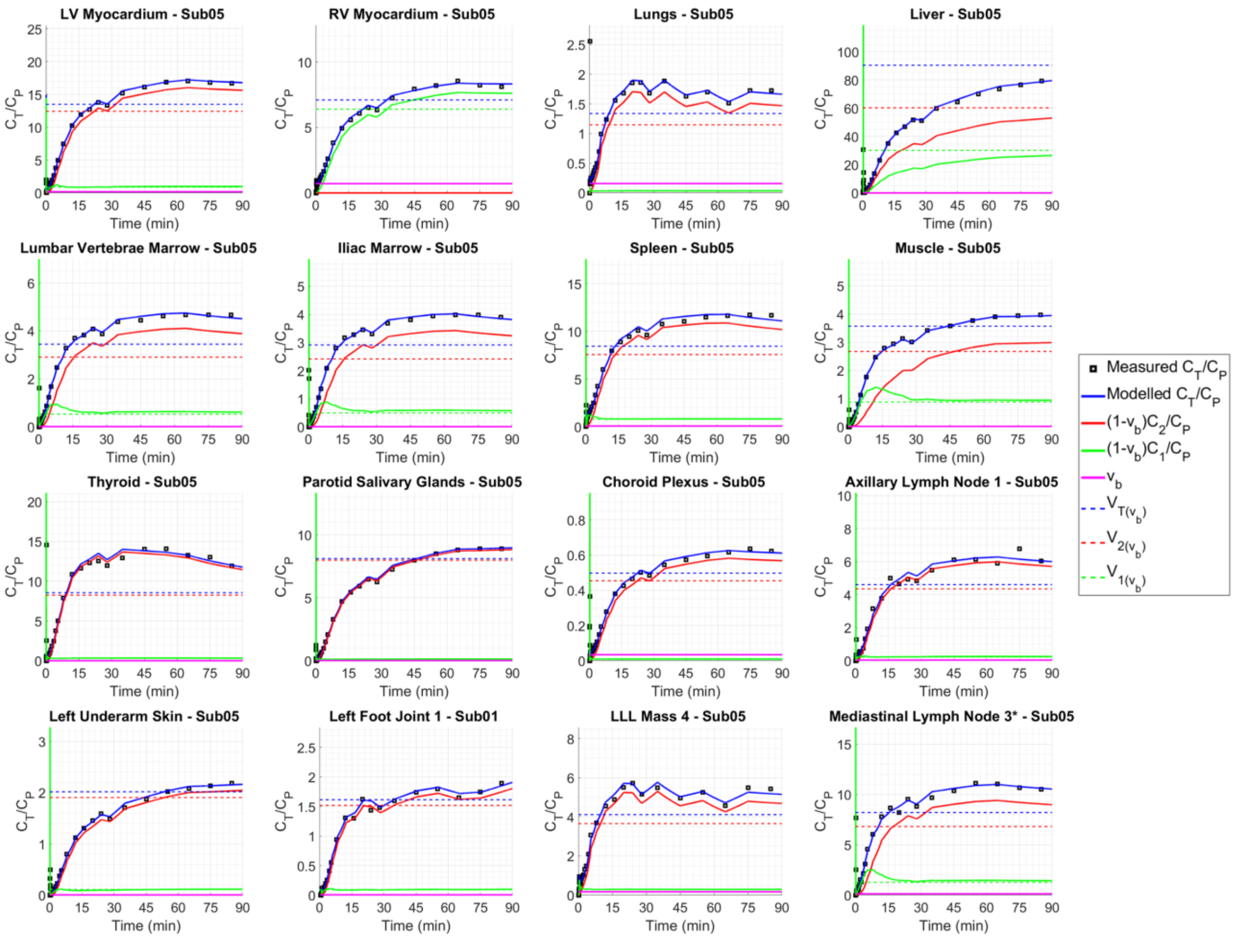


**SUPPLEMENTAL FIGURE 20.** Examples of SUVR curves (black squares) in different organs-of-interest compared to modeled SUVRs (solid blue) and blood-volume corrected total volume of distribution (dashed blue). The blood-volume corrected SUVRs of the first (solid red) and the second (solid green) compartments are additionally compared to blood-volume corrected volumes of distribution in the (dashed red) and the second (dashed green) compartments. The original measured blood-pool TACs were used for calculation of SUVRs in all cases.

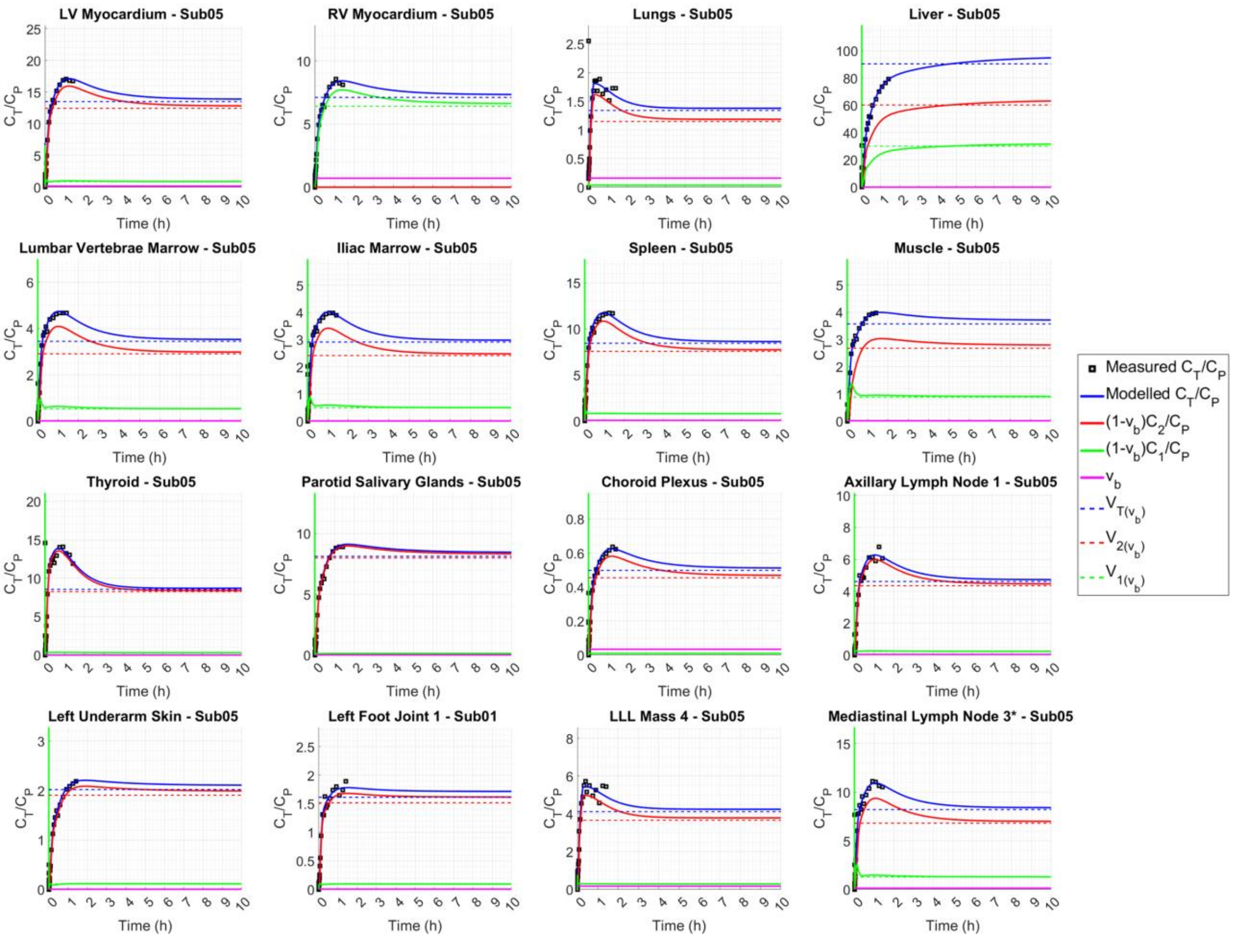


**SUPPLEMENTAL FIGURE 21.** Examples of SUVR curves (black squares) extrapolated to 10 h p.i. in different organs-of-interest compared to modeled SUVRs (solid blue) and blood-volume corrected total volume of distribution (dashed blue). The blood-volume corrected SUVRs of the first (solid red) and the second (solid green) compartments are additionally compared to blood-volume corrected volumes of distribution in the (dashed red) and the second (dashed green) compartments. The extrapolations of the biexponentially fitted blood-pool TACs were used for calculation of SUVRs in all cases.

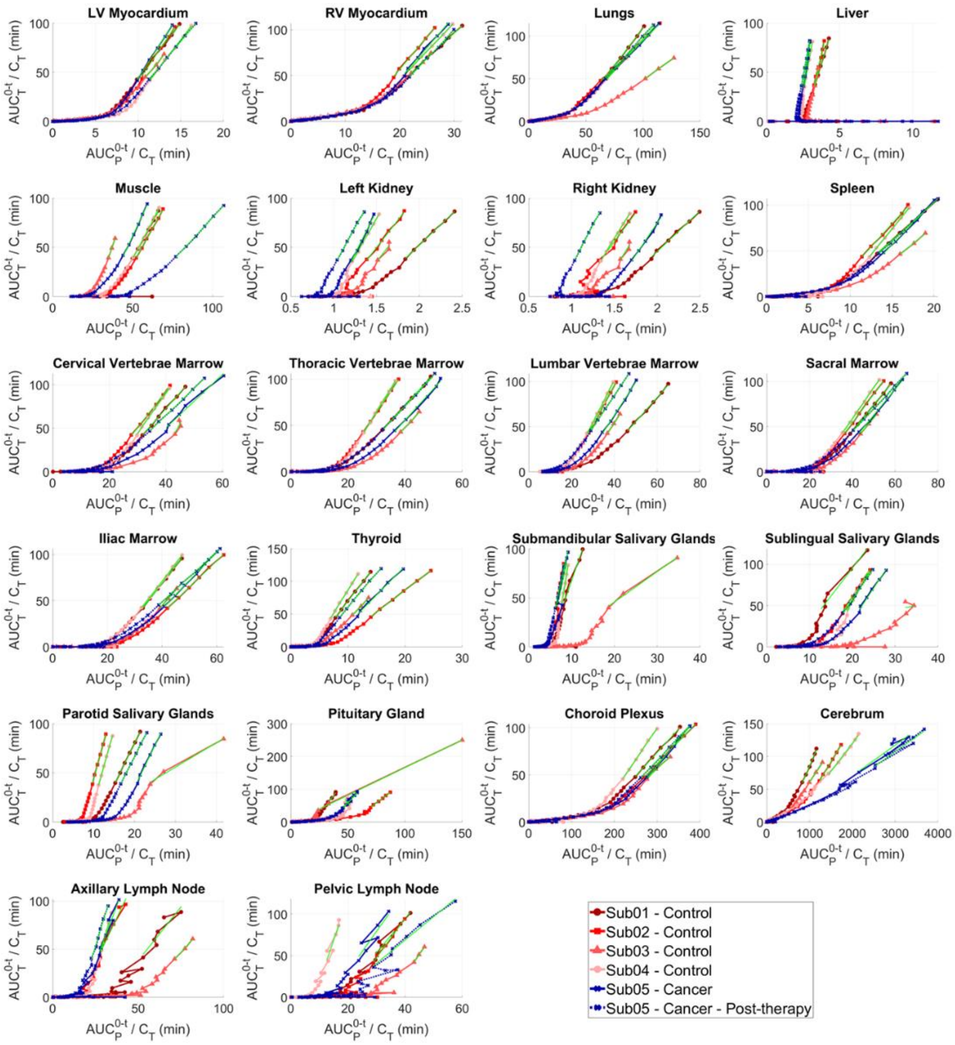


**SUPPLEMENTAL FIGURE 22.** Logan plots of healthy organs-of-interest analyzed in all subjects fitted with t*=30 min. The VOIs in the head region of Sub03 were affected by subject's head motion.

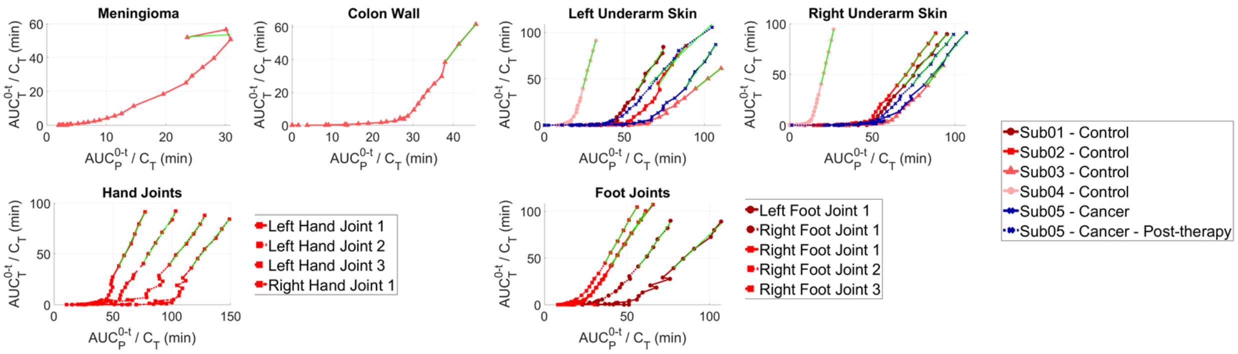


**SUPPLEMENTAL FIGURE 23.** Logan plots of organs with focally high uptake pattern in all subjects fitted with t*=30 min. The VOIs in the head region of Sub03 were affected by subject's head motion.

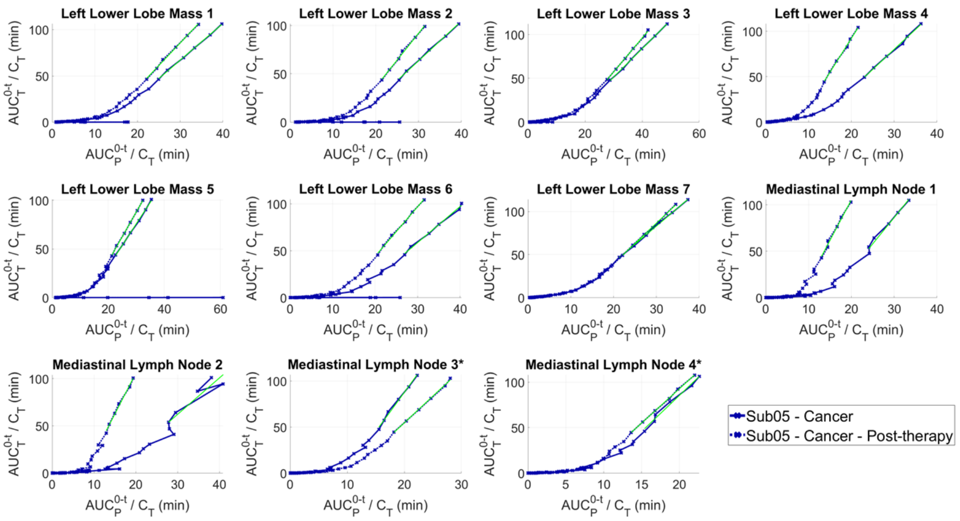


**SUPPLEMENTAL FIGURE 24.** Logan plots of seven sub-regions of the LLL mass and the four mediastinal LNs of Sub05 pre- and post-therapy fitted with t*=30 min.

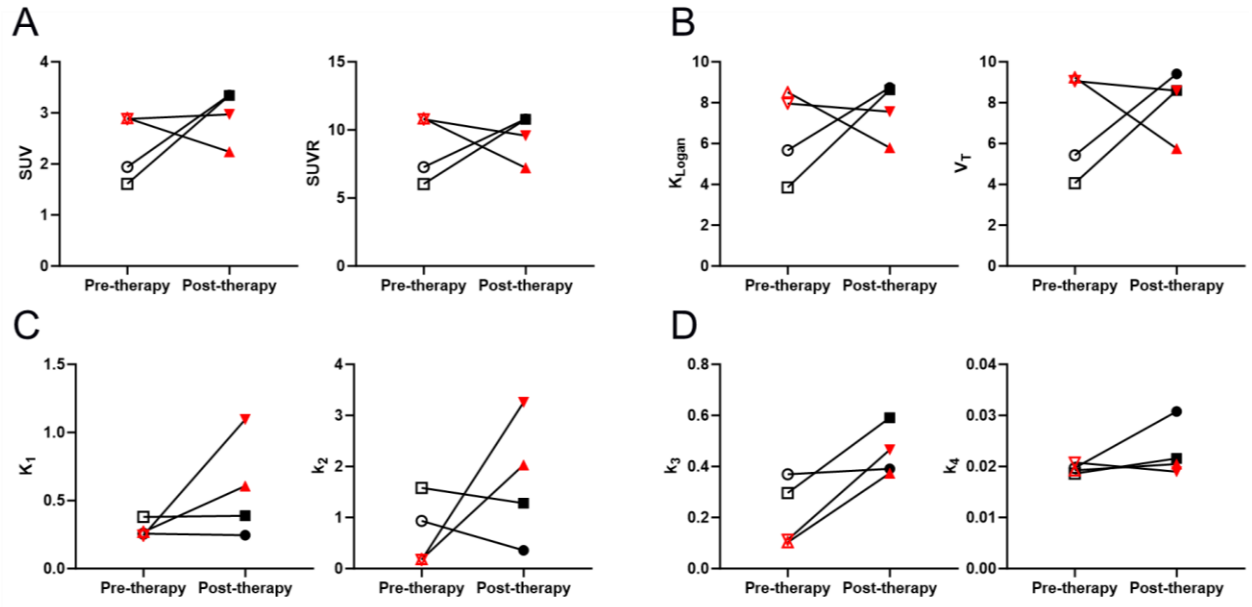


**SUPPLEMENTAL FIGURE 25.** Changes in non-enlarged (black) and enlarged (red) mediastinal LNs pre-therapy and post-therapy, using (A) SUVpeak or SUVR values without kinetic modeling, (B) $\boldsymbol{K_{Logan}}$ from Logan plots or $\boldsymbol{V_T}$ from 2T6P compartmental modeling as macroparameters with high identifiability, (C) 2T6P rate constants between blood and first compartment ($\boldsymbol{K_1}$and $\boldsymbol{k_2}$), and (D) 2T6P rate constants between first and second compartments ($\boldsymbol{k_3}$and $\boldsymbol{k_4}$).

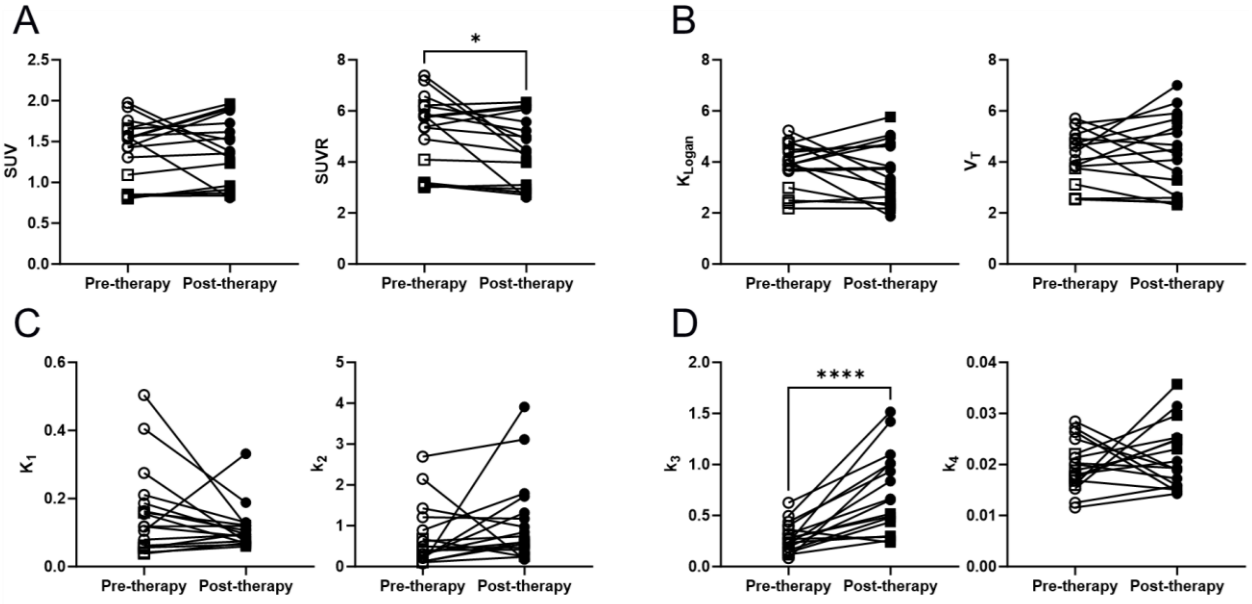


**SUPPLEMENTAL FIGURE 26.** Changes in axillary (circles) and pelvic (squares) LNs pre-therapy and post-therapy, using (A) SUVpeak or SUVR values without kinetic modeling, (B) $\boldsymbol{K_{Logan}}$ from Logan plots or $\boldsymbol{V_T}$ from 2T6P compartmental modeling as macroparameters with high identifiability, (C) 2T6P rate constants between blood and first compartment ($\boldsymbol{K_1}$and $\boldsymbol{k_2}$), and (D) 2T6P rate constants between first and second compartments ($\boldsymbol{k_3}$and $\boldsymbol{k_4}$).
